# FINEMAP-miss: Fine-mapping genome-wide association studies with missing genotype information

**DOI:** 10.1101/2025.09.30.25336878

**Authors:** Joonas Kartau, Matti Pirinen

## Abstract

**Motivation:** The most informative genome-wide association studies (GWAS) are meta-analyses that have combined multiple studies to increase the GWAS sample size. Statistical fine-mapping is a key downstream analysis of GWAS to jointly evaluate the probability of causality of all variants in a genomic region of interest. Current fine-mapping methods are miscalibrated in the meta-analysis setting due to variation in sample size across the variants.

**Results:** We introduce FINEMAP-miss, a new fine-mapping method that extends the FINEMAP model to account for variant-specific missingness. We show that FINEMAP-miss is well-calibrated in meta-analysis simulations where the standard fine-mapping fails. Compared to the summary statistics imputation approach, FINEMAP-miss provides clear improvement when the causal variants have low imputation information or when the sample size or complexity of the meta-analysis setting increase. We successfully apply FINEMAP-miss on a breast cancer GWAS meta-analysis where neither the standard fine-mapping nor the summary statistics imputation are applicable.

**Availability:** An open source implementation of FINEMAP-miss as an R package (“finemapmiss”) is available at https://github.com/JoonasKartau/finemapmiss.

## 1 Introduction

Genome-wide association study (GWAS) is a central tool to map genomic regions to complex phenotypes. A standard GWAS measures marginal associations of variants and ignores the extensive correlation structure between the variants caused by linkage disequilibrium (LD). Therefore, it is a common practice to refine the GWAS results by fine-mapping methods, such as CAVIAR [1], FINEMAP [2] or SuSiE [3], that jointly model the GWAS summary statistics of multiple variants and their LD to provide probabilistic inference on the causal variants behind the associations observed.

The largest and most informative GWAS tend to be meta-analyses that combine information across multiple biobanks and other studies. The sample sizes of the meta-analyzed variants often vary because every variant has not been present in every study, for example, due to the use of different genotyping chips, imputation panels or quality control methods. Since the current fine-mapping methods do not account for variation in information content between the variants, they are often miscalibrated in the meta-analysis setting [4].

Previous approaches to fine-mapping meta-analysis data have suggested a complete removal of the variants that either show inconsistency with their neighbors (SLALOM [4] and CARMA [5]) or that are not observed in every cohort [6]. Both of these approaches have the downside that they may remove causal variants from the analysis, still leading to miscalibrated results.

In this work, we introduce FINEMAP-miss, a new fine-mapping method that extends the FINEMAP model [2] to account for incomplete sample overlap between variants. In particular, FINEMAP-miss allows all variants that have any data in a GWAS meta-analysis to be included in the fine-mapping. We compare the performance of FINEMAP-miss to summary statistics imputation [7, 8] and apply it to a GWAS meta-analysis of breast cancer [9].

## 2 Methods

### 2.1 FINEMAP model

Here, we summarize the FINEMAP model [2], which is the starting point for our work.

Let ***y*** ∈ ℝ^*n*^ be a quantitative phenotype vector for a sample of size *n*, and ***X*** ∈ ℝ^*n*×*p*^ a matrix of genotypes for *p* variants, where ***y*** and each column of ***X*** is centered and standardized to have a variance of 1. We assume the following linear model between the genotypes and the phenotype:

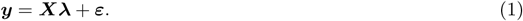

Here, individual-specific errors *ε*_*i*_ ∼ 𝒩 (0, 1 − ***λ***^⊺^***Rλ***) are independent and normally distributed for *i* ≤ *n* and 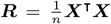 is the correlation matrix of the variants. In the fine-mapping context, ***R*** is called the linkage disequilibrium (LD) matrix and we expect that only *k* ≪ *p* variants have non-zero effects; hence ***λ*** is a sparse vector with *p* − *k* zeros. We assume that the non-zero elements of ***λ*** have a prior distribution 𝒩 (0, *τ*^2^), where *τ*^2^ is a fixed parameter. Let ***γ*** ∈ {0, 1}^*p*^ denote a causal configuration with *γ*_*j*_ = 1 if and only if *λ*_*j*_ ≠ 0. The model is completed by a prior distribution *p*(***γ***) on the set of all possible causal configurations. The default choice in the FINEMAP software is a truncated binomial distribution

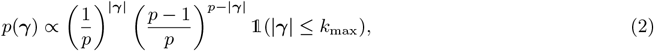

where |***γ***| is the number of non-zero elements of ***γ***, *k*_max_ is the fixed upper bound of the non-zero effects allowed by the model and is 𝟙 the indicator function.

We assume no access to the full data (***y, X***) but only to GWAS results consisting of estimates of the marginal effects 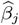 from univariate linear models ***y*** = ***x***_*·,j*_ *β*_*j*_ + ***ε***_*j*_, together with the corresponding standard errors *s*_*j*_ and the LD matrix ***R***. (See Supplementary Data for details of GWAS results.) To compare how the data support each causal configuration ***γ***, FINEMAP uses the marginal likelihood

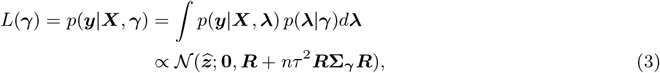

where 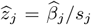 for all *j* ≤ *p* and **Σ**_***γ***_ = Diag(***γ***). Thus, *L*(***γ***) depends on the data (***y, X***) only through the marginal z-scores 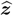 and the LD matrix ***R***.

FINEMAP is efficient because the Bayes factor BF_***γ***_ = *L*(***γ***)*/L*(***γ***_0_), comparing ***γ*** to the null configuration ***γ***_0_ = (0, …, 0), can be computed very quickly. Denote by *C* = {*j*: *γ*_*j*_ = 1} the set of causal variants of ***γ*** and by *N* its complement set of the non-causal variants. Then

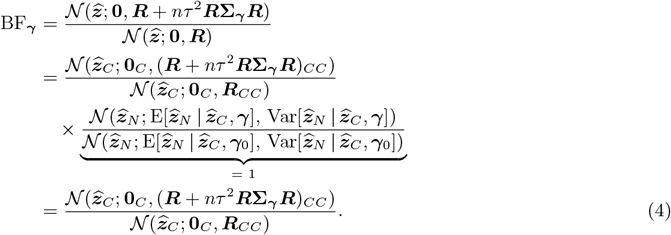

As the contribution of the non-causal variants cancels out from BF_***γ***_, we are left with evaluating only |***γ***|-dimensional densities rather than *p*-dimensional densities. FINEMAP uses a Shotgun Stochastic Search algorithm [10] to evaluate relevant configurations.

### 2.2 Genotype imputation info scores

The original FINEMAP model was specified for the standardized genotypes. However, typically GWAS software outputs the effects per allele, and for the imputed variants [11], the genotype dosage (expected value of the genotype based on the imputation) is used as the predictor in the regression model. We can transform the allelic effects to the scale of standardized genotypes by multiplication with 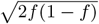, where *f* is the minor allele frequency of the variant. The variance of the corresponding scaled genotype dosage of variant *j* is Var 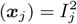, where 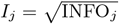 is the square root of the imputation info score of variant *j* defined in Supplementary Data. When the genotype is observed, *I*_*j*_ = 1, but when it is imputed, *I*_*j*_ is typically below 1 as some information has been lost in the imputation.

In Supplementary Data, we show that the imperfect INFO scores modify two key relationships of the FINEMAP model. First, the LD matrix ***R***, that we define as the correlation matrix of the predictors, is now 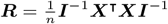 where ***I*** = diag(*I*_*j*_). Second, the relationship between the causal and marginal effects becomes 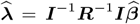. Both of these relationship revert back to the original FINEMAP model if 𝕀 = I_*p*_ (identity matrix of dimension *p*).

### 2.3 Covariance matrix with missing information

Variant-specific missingness in GWAS results can be presented in the inverse-variance weighted meta-analysis framework. Assume that the meta-analysis results at variant *j* were generated by combining GWAS results 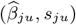 from *u* = 1, …, *d* independent studies using the meta-analysis framework (Supplementary Data). Thus, the meta-analyzed z-scores are

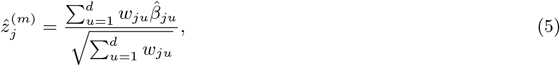

where the weights 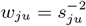. To account for missing data we set *w*_*ju*_ = 0 if variant *j* was not observed in study *u*. Both the correlation and the covariance between the meta-analyzed z-scores of variants *j*_1_ and *j*_2_ is

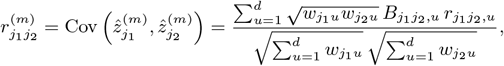

Where 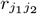,*u* is the correlation between 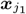 and 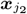 in study *u* and

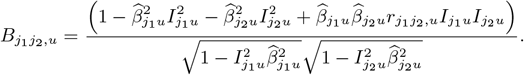

We denote the covariance matrix of the meta-analyzed z-scores by

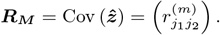

### 2.4 FINEMAP-miss

In a meta-analysis with missing data, we show in Supplementary Data that the marginal likelihood of ***γ*** takes the form

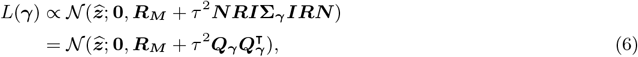

where ***N*** is a *p* × *p* diagonal matrix containing the square root of the variant-specific sample sizes, and we have defined a (*p* × |***γ***|)-matrix

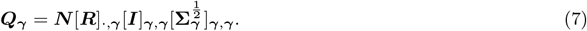

The logarithm of the Bayes factor of ***γ*** becomes

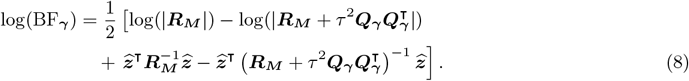

Using the Woodbury matrix identity and the matrix determinant lemma, the inverse matrix 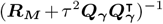 and the corresponding determinant 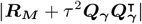 can be written as

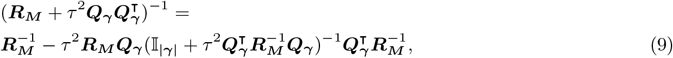

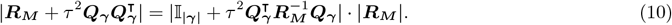

Importantly, the matrix 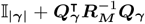 is of dimension |***γ***| × |***γ***|, and (8) takes a simpler form without any large matrices that depend on ***γ***:

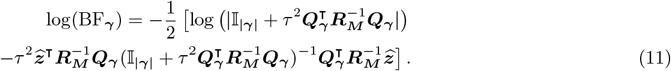

Thus, the expensive computation of an inverse and a determinant of a *p* × *p* matrix is required only once for ***R***_***M***_ during the whole algorithm and the subsequent operations can be done with low dimensional matrices. In Supplementary Data, we give the technical details of an efficient and robust implementation of these computations in our software package for FINEMAP-miss.

### 2.5 Summary statistics imputation

As an alternative to FINEMAP-miss, we could try to impute the missing GWAS results before the meta-analysis, separately in each study, using summary statistics imputation [7, 8].

Within one study, let *O* denote the subset of observed z-scores, and *j* be one unobserved variant. The joint distribution of the marginal z-scores under the null model is

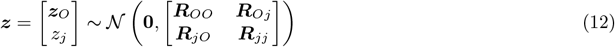

The unobserved z-score can be imputed given the observed z-scores under the null model as

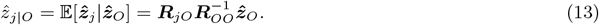

These imputed z-scores are diluted due to a loss of information and a rescaling by the standard error of the imputed z-score has been proposed [7]:

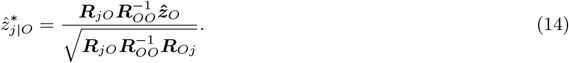

Given either 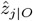 (“unscaled imputation”) or 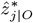 (“scaled imputation”), it is possible to perform a meta-analysis and run FINEMAP as if we had no missing data. However, in the meta-analysis setting with missing data, accurate summary statistics imputation requires access to the original study-specific GWAS results which are not always available. In contrast, with FINEMAP-miss the meta-analyzed results alone are sufficient as long as we also know which study contributed data to which variant.

### 2.6 Quantities to evaluate fine-mapping

**Posterior inclusion probability (PIP)** for variant *j* describes the probability that *j* is one of the causal variants:

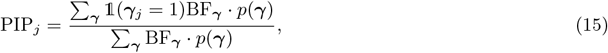

where *p*(***γ***) is the prior probability of the configuration ***γ***. By default, we use the truncated binomial prior (2).

#### PIP bins

Following [12], we can evaluate the accuracy of the PIPs by stratifying the variants into bins based on their estimated PIP and examining the proportion of causal variants in each bin. If the fine-mapping model is calibrated correctly, then the proportion of causal variants should be close to the mean of the PIPs of the variants in the bin (details in Supplementary Data).

#### Posterior expected number of causal variants (PENC)

The FINEMAP model provides the posterior distribution for the number of causal variants *K*

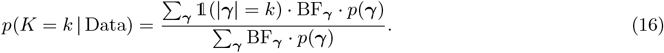

The PENC is the expectation of this distribution [6]:

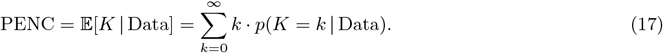

#### Credible set (CS)

The aim of a CS is to collect together a (small) set of variants that includes a causal variant with a specified probability *θ*. For a fixed number of causal variants *k*, we construct CSs as follows.

1. Identify the top configuration with *k* causal variants ***γ***_*k*_.
2. For each causal variant *j* ∈ ***γ***_*k*_,
  a. Compute vector ***b*** of BFs for configurations (***γ***_*k*_ − ***e***_*j*_ + ***e***_*l*_) where *l* ∈ ({1, …, *p*} *\* ***γ***_*k*_) ∪ {*j*} and ***e***_*j*_ is the indicator vector for position *j*.
  b. Normalize to obtain conditional posterior probability vector 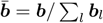.
  c. Select variants in decreasing order based on 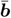 until the sum of probabilities exceeds *θ*.

**Credible set power** is defined as the probability that a causal variant is included in a CS reported by FINEMAP or FINEMAP-miss.

## 3 Data

### 3.1 Data generation

The data for the simulations were generated using the imputed genotype data from UK biobank (UKB) [13]. To ensure homogeneous sample, we considered *n* = 300, 000 individuals from “white, British” subset with no close relatives included. We considered chromosme 2 region 26,688,494-27,088,494 (coordinates in build 37) and filtered out variants with MAF ≤ 0.1% or INFO score ≤ 0.1, leaving *p* = 1925 variants for the simulations. This filtering was performed using PLINK v1.9 [14].

To incorporate the uncertainty arising from genotype imputation, a set of “virtual” genotypes were sampled for each individual *i* and variant *j* using the available genotype imputation probabilities (*p*_*ij,g*_) for genotype *g* ∈ {0, 1, 2} similarly as [12]. Let ***G***^(*v*)^ ∈ ℝ ^*n*×*p*^ denote the virtual genotype matrix, whose elements 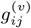 are realizations of random variables 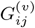 with the following probability mass function:

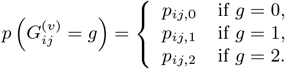

We denote by ***X***^(*v*)^ the matrix of scaled virtual genotypes that resulted when the columns of ***G***^(*v*)^ were standardized. The phenotype was generated using the scaled genotypes:

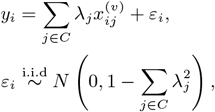

where *C* is the set of causal variants and the effect sizes *λ*_*j*_ were equal and jointly explained at most 0.1% of the phenotypic variance. We considered settings with one or two causal variants.

We generated two types of simulations depending on the INFO of the causal variant. In the high INFO simulations the causal variants were restricted to have INFO ≥ 0.9, and in the low INFO simulations, INFO *<* 0.9. In the latter simulations, the causal variant INFO scores ranged in [0.352, 0.900). In each simulation type, the causal variants *j* were sampled uniformly at random from the set of all variants that met the given INFO score criterion. For conducting the GWAS, the genotype dosages (rather than virtual genotypes) were used, to model a realistic GWAS.

The data were randomly split into three subcohorts, each containing 100,000 individuals. A linear model GWAS were performed independently on the first two cohorts, resulting in two sets of GWAS results. The third cohort was used as an LD reference in fine-mapping. Generation of phenotype data, linear model GWAS, and LD matrices were performed using Rbgen v1.1.5 [15] and vcfR v1.15.0 [16].

At each variant, data were missing from exactly one, randomly chosen GWAS cohort with a probability of *p*_*m*_ = 0.2. The causal variants in each simulation were observed in only one GWAS cohort. The GWAS results of the two subcohorts were meta-analyzed. Fine-mapping was performed on the meta-analyzed data with FINEMAP v.1.4.2 (www.finemap.me) with and without summary statistics imputation, and with FINEMAP-miss v.1.0.

### 3.2 Large sample meta-analysis simulations

To investigate the effect of increasing the sample size up to 1,000,000, and the number of meta-analyzed data sets up to 10, we created 8 additional data sets, each by sampling with replacement 100,000 individuals from the first subcohort from section 3.1.

For the resampled subcohorts, we generated phenotypes using the same causal variants as in section 3.1. Here the possible number of subcohorts for missing variants ranged from 1 to 9, with equal probability. The causal variants in each simulation were observed in only one subcohort.

### 3.3 Computation time

To evaluate the run time, we generated GWAS summary statistics from a multivariate normal distribution for an increasing number of variants between 1,000 and 20,000 with 5 simulations per each count.

Each simulation contained one causal variant that explained 0.1% of the phenotypic variance. For both FINEMAP-miss v.1.0 and FINEMAP v1.4.2, exactly 50 Shotgun Stochastic Search neighborhoods were evaluated.

We used FINEMAP-miss v.1.0 run in R v.4.0.0 and FINEMAP v.1.4.2, both run on a virtual machine with 16 CPUs (Intel Xeon - 2.20 GHz) and 128 GB of memory.

### 3.4 Breast cancer meta-analysis

We performed fine-mapping of a breast cancer meta-analysis by Rashkin et al [9]. Data were downloaded from the NHGRI-EBI GWAS Catalog [17] on 03.03.2025 (study GCST90011804). This meta-analysis combined breast cancer GWAS from UKB and from the Kaiser Permanente Genetic Epidemiology Research on Adult Health and Aging cohort (GERA). Effective sample sizes (computed as *nϕ*(1 − *ϕ*) with *n* as the total sample size and *ϕ* the proportion of cases) for the studies were 12,954 and 3,510, respectively.

The region was centered at the top signal on chromosome 10 near the *FGFR2* gene, and contained common (MAF *>* 1%) variants extending 1 Mb to either side (10:122340781-124340113). As the reference LD panel we used the full set of “white, British” individuals from UKB (*n* = 327, 307).

The data contained only the meta-analyzed odds-ratios and p-values but no study-specific summary statistics. Therefore, it was not possible to perform accurate summary statistics imputation. We still attempted fine-mapping after approximate summary statistics imputation that assumed that when a variant had been observed in both studies, then its effect size was equal between the studies. (Supplementary Data.)

Since FINEMAP and FINEMAP-miss require an LD reference, some variants that were present only in GERA had to be omitted, as the LD reference available to us was from UKB.

## 4 Results

We report the simulations with one causal variant (high and low INFO) here and the simulations with two causal variants in Supplementary Data.

We observed that FINEMAP without imputed summary statistics is severely miscalibrated in meta-analyses with missing information. From low credible set power and large maximal non-causal PIPs (MNC-PIP) in Table 1, we conclude that it lacks both sensitivity and specificity for detecting causal variants and cannot be trusted in meta-analyses with missing data. We also observed that the scaled imputation performed better than the unscaled imputation in all simulation scenarios. Therefore, we next focus on comparing FINEMAP-miss to FINEMAP with scaled imputation.

**Table 1:** Means of credible set power (CS power), PIP of causal variant (C-PIP), maximal PIP of non-causal variants (MNC-PIP) and posterior expected number of causal variants (PENC) from the simulations with one causal variant, separated by whether the causal variant has high INFO (≥ 0.90) or low INFO (*<* 0.90) and by sample sizes *n* = 2e+5 or *n* = 1e+6. FINEMAP v.1.4.2 (FM) was run with three versions of the input data: scaled imputation, unscaled imputation or no imputation. imp. = imputed data.

|  |  | Method |  |  |  |
| --- | --- | --- | --- | --- | --- |
|  |  | FINEMAP-miss | FM Scaled imp. | FM Unscaled imp. | FM No imp. |
| $n = 2e+5$ | | | | | |
| High INFO | CS power | 0.97 | 0.95 | 0.95 | 0.87 |
|  | C-PIP | 0.500 | 0.511 | 0.478 | 0.106 |
|  | MNC-PIP | 0.175 | 0.167 | 0.191 | 0.938 |
|  | PENC | 1.137 | 1.144 | 1.133 | 4.148 |
| Low INFO | CS power | 1.00 | 1.00 | 1.00 | 0.78 |
|  | C-PIP | 0.948 | 0.943 | 0.842 | 0.591 |
|  | MNC-PIP | 0.056 | 0.051 | 0.122 | 0.627 |
|  | PENC | 1.129 | 1.125 | 1.119 | 2.460 |
| $n = 1e+6$ | | | | | |
| High INFO | CS power | 0.98 | 0.99 | 0.88 | 0.40 |
|  | C-PIP | 0.619 | 0.628 | 0.490 | 0.400 |
|  | MNC-PIP | 0.162 | 0.234 | 0.346 | 1.000 |
|  | PENC | 1.080 | 1.255 | 1.282 | 4.964 |
| Low INFO | CS power | 1.00 | 0.99 | 0.93 | 0.73 |
|  | C-PIP | 0.963 | 0.964 | 0.707 | 0.574 |
|  | MNC-PIP | 0.040 | 0.409 | 0.752 | 0.966 |
|  | PENC | 1.070 | 1.584 | 1.917 | 4.208 |

For the smaller sample size *n* = 2e+5, FINEMAP-miss and FINEMAP with scaled imputation performed well, as each method displayed high credible set power (≥ 0.95), high mean PIPs of the causal variants (C-PIP), while also keeping MNC-PIPs low.

With both methods, the C-PIP and MNC-PIP values differ depending on whether the causal variant has high or low INFO. This is likely because the variants with high INFO tend to be more strongly correlated with other variants than the variants with low INFO. Thus, there are more non-causal variants that tag the causal signal in the high INFO scenario, leading to lower C-PIPs and higher MNC-PIPs compared to the low INFO scenario. FINEMAP-miss and FINEMAP with imputed summary statistics all provided similar PENC values.

For the smaller sample size, the PIP bins in Figure 1 do not indicate miscalibration for FINEMAP-miss or the scaled imputation, but the unscaled imputation is not calibrated well for the highest bin of [0.9,1].

**Figure 1:**
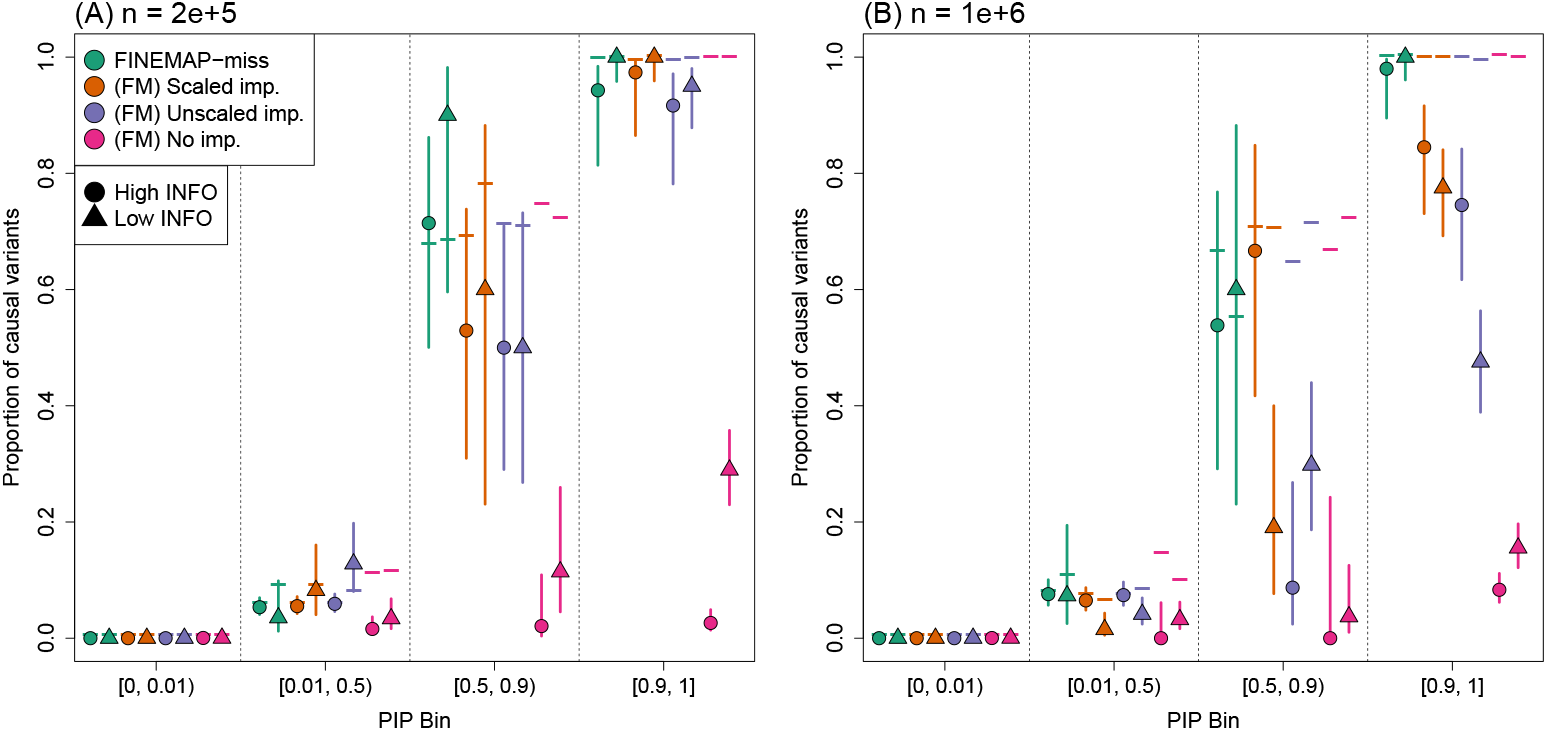
PIP bin calibration plot from high INFO (circles) and low INFO (triangles) simulations. Symbols indicate the observed proportion of causal variants. Horizontal lines indicate the expectation given by the average PIPs. 95% Wilson confidence intervals are provided as vertical lines. The simulation sample size is 2e+5 in panel A and *n* = 1e+6 in panel B. FM = FINEMAP v.1.4.2. imp. = imputed data.

When the sample size and the number of datasets in the meta-analysis increased (*n* = 1e+6), there was an increase in credible set power and C-PIPs. A clear difference between the methods also appeared in the MNC-PIP values. For the scaled imputation, MNC-PIP increased indicating less accurate fine-mapping compared to the smaller sample size, especially in the low INFO simulations. In contrast, for FINEMAP-miss, MNC-PIP decreased indicating that the accuracy of FINEMAP-miss improved with the sample size (Table 1).

For the scaled imputation, 10% of the high INFO simulations contained a non-causal variant with PIP *>* 0.8 (Figure 2). The respective value in the low INFO simulations was 30%. This enrichment of high PIP non-causal variants is also visible in Figure 1, where the highest bin is miscalibrated for both modes of imputation. We observed much lower summary statistics imputation quality for the causal variants in the low INFO simulations, which we believe to be the driving force of the miscalibration of fine-mapping with imputed summary data.

**Figure 2:**
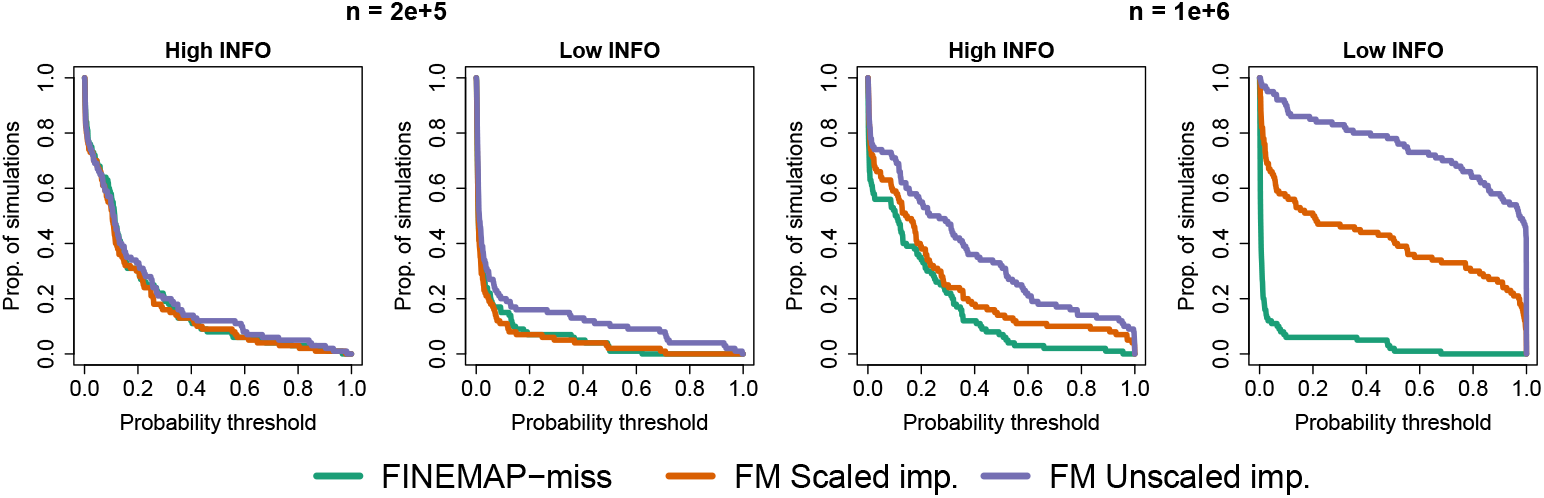
Proportion of simulations (y-axis), whose fine-mapping results contained a non-causal variant with a PIP above a probability threshold (x-axis). Results displayed for simulations with sample sizes *n* = 2e+5 and *n* = 1e+6, and for both high and low INFO causal variant simulations. FM = FINEMAP v.1.4.2. imp. = imputed data.

No signs of miscalibration were observed with FINEMAP-miss. In only 2% of the high INFO and none of the low INFO simulations was there a non-causal variant with a PIP *>* 0.8, and the PIP bins remained well calibrated across the scenarios.

### 4.1 Computation Time

Due to the matrix operations, the time complexity of FINEMAP-miss is in the order of 𝒪 (*p*^3^), where *p* is the number of variants, while for FINEMAP the complexity is growing approximately linearly in *p*. In practice, FINEMAP-miss completes an analysis with *p* = 20, 000 variants in under 19 minutes using standard hardware (specified in Section 3.3), keeping typical fine-mapping analyses possible to run with FINEMAP-miss. (Supplementary Figure 9.)

### 4.2 Breast cancer meta-analysis

FINEMAP-miss identified two signals, whose credible sets gave the highest posterior probability of being causal to rs9421410 and rs2912780, respectively.

When comparing the methods (Fig. 3), we observe that FINEMAP-miss (B) lists the lead SNPs and the variants highly correlated with them as plausible candidates, displaying the expected fine-mapping behavior. Since we had no access to the study-wise effect estimates, we had to rely on an approximate imputation approach that assumed that the effect size estimates were equal between the studies. With such an approximate version of scaled imputation (C), FINEMAP captured the same two signals as FINEMAP-miss, however, with an apparent bias where variants with missing information received higher PIPs than their highly correlated neighbours. For example, the variants rs9421409 and rs9421410 are perfectly correlated (*r*^2^ = 1) across the 1000 Genomes populations [18], but here they misleadingly got very different PIPs after the approximate imputation because only one of them (rs9421410) was present in both studies of the meta-analysis. (The results from unscaled imputation were nearly identical to scaled imputation and are not shown here.) FINEMAP without imputed summary statistics (D) gave PIPs close to one for multiple variants, indicating severe model miscalibration.

**Figure 3:**
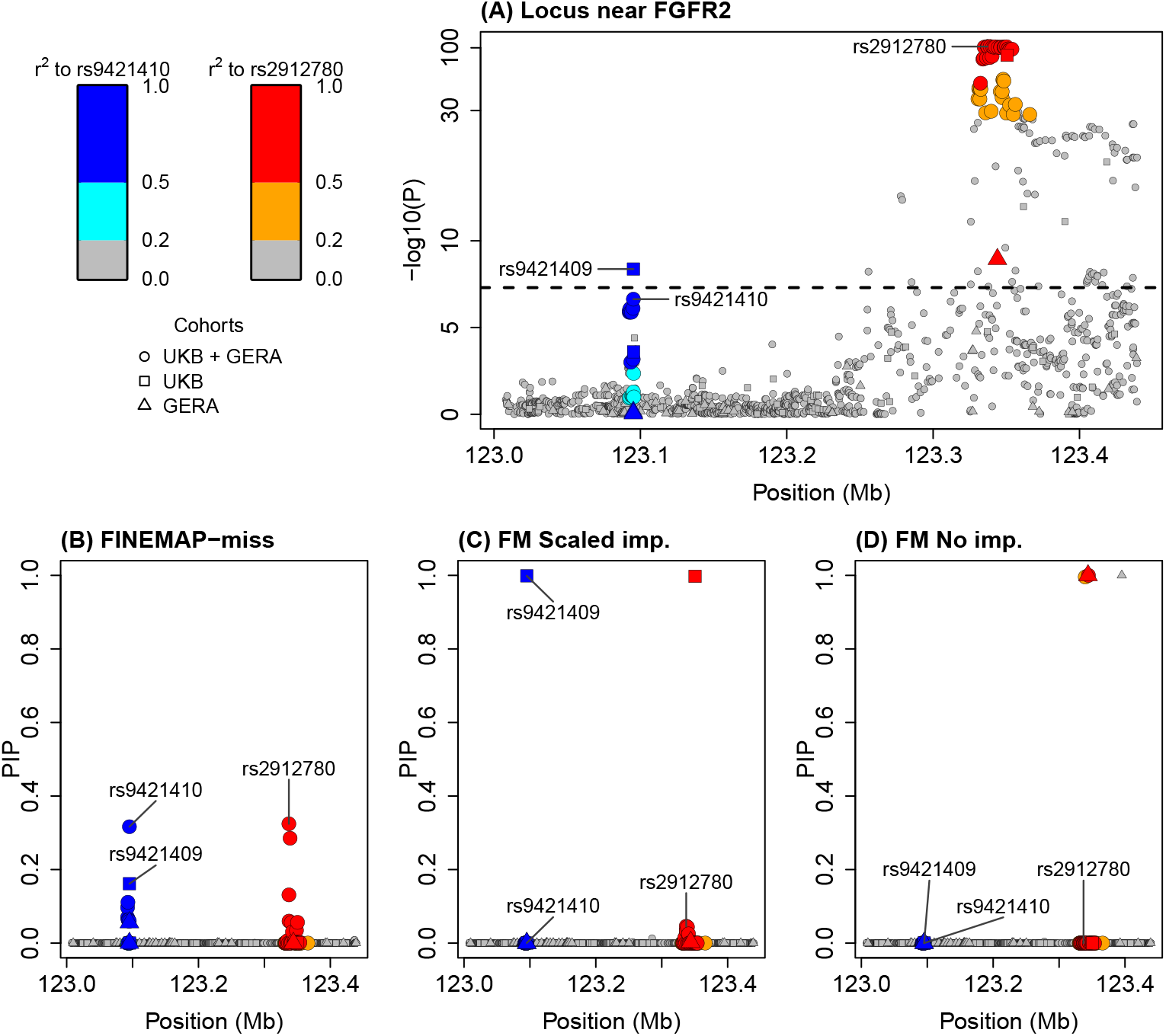
Breast cancer meta-analysis with two cohorts (UKB + GERA) for the *FGFR2* locus. (A) Marginal p-values, posterior inclusion probabilitiy (PIP) from fine-mapping with (B) FINEMAP-miss, (C) FINEMAP with approximate scaled imputation, and (D) FINEMAP without imputation. Three variants mentioned in the text are named. Symbols describe in which cohorts the variant was present. Colors denote *r*^2^ to rs9421410 (blue scale) or rs2912780 (red scale) according to the color legend. The scale of the y-axis in plot (A) is non-linear. FM = FINEMAP v.1.4.2. imp. = imputed data.

To understand the miscalibration of FINEMAP with approximate summary statistic imputation, we note that the meta-analysis results report the effect estimate of rs9421410 to be highly heterogenous (heterogeneity index *I*^2^ = 0.87) between UKB and GERA. However, as the meta-analysis results do not include the study-wise effect estimates of rs9421410, the approximate imputation wrongly assumes that the effect sizes are equal between the studies. Here, this leads to an overestimated imputed effect size for rs9421409.

## 5 Discussion

Meta-analyses of GWAS data sets provide a way to increase statistical power to detect causal genetic variants but missing data complicates fine-mapping of meta-analyzed data with standard methods [1, 2, 3]. We have developed FINEMAP-miss that models the missingness in GWAS meta-analyses and can provide accurate fine-mapping even in cases where the standard fine-mapping methods fail.

An alternative approach to fine-map meta-analysis data is based on strict quality control filtering where variants that indicate discrepancies between the association statistics and the LD structure are removed from the analysis [4, 5, 6]. While this approach leads to consistent results on the remaining variants, it risks removing causal variants from the analysis, and therefore may lead to biased results.

Another approach to fine-map meta-analysis data is to impute the missing GWAS summary statistics in each data set [7, 8] before the meta-analysis and run a standard fine-mapping on the meta-analyzed data. This approach works well when the data on the true causal variants is of high quality but, based on our results, starts to produce false positives at much higher rate than FINEMAP-miss as the total GWAS sample size increases and/or the imputation information at the causal variants decreases. Summary statistics imputation also requires access to the cohort specific effect size estimates, but publicly available GWAS meta-analyses typically provide only the combined effect estimates. For such data, accurate imputation is not possible using the existing imputation methods, as we demonstrated with the breast cancer meta-analysis example.

While FINEMAP-miss extends our possibilities to fine-map GWAS meta-analyses, there still remain limitations related to the applicability of FINEMAP-miss. First, FINEMAP-miss requires the exact study-wise missingness structure to accurately fine-map a meta-analysis. Luckily, this can often be inferred from the meta-analysis data, if the total sample size of each meta-analyzed variant and the sample size of each dataset is known. While FINEMAP-miss is slower than standard fine-mapping software packages, such as FINEMAP and SuSiE, we have demonstrated that FINEMAP-miss is still practical to run for typical regions that contain up to 10,000 variants. The most serious issue that will continue to hamper meta-analysis fine-mapping also with FINEMAP-miss is the possibility that accurate LD is not available. For example, in a migraine meta-analysis combining Finnish and non-Finnish, European GWAS, a use of weighted LD reference from FinnGen and UK biobank provided accurate fine-mapping results (compared to the in-sample LD) in 24 out of 26 regions but in the two remaining regions the fine-mapping based on the LD reference failed [6]. Additionally, FINEMAP-miss requires that the reference LD matrix includes also the partially missing variants. Hence, we would need to have access to a high-quality LD reference panel from suitable genetic ancestry that contains information on all variants included in the meta-analysis. Topics for future study include how to detect when fine-mapping with reference LD has been reliable and how to adjust the fine-mapping models to account for the level of accuracy of the reference LD.

Recent development of fine-mapping methods include cross-population approaches such as XMAP [19], MESuSiE [20] and SuSiEx [21]. These methods provide additional power to detect causal variants by leveraging the unique LD structures present in different ancestries. In addition, it has been shown that incorporation of infinitesimal effects in the fine-mapping model (FINEMAP-inf, SuSiE-inf) [12], and the inclusion of functional annotations as prior information, such as with PAINTOR [22], PolyFUN + FINEMAP/SuSiE [23], or CARMA [5], improves fine-mapping. In the future, combining the useful properties of these approaches with the missingness model of FINEMAP-miss could help us to extract additional information from existing GWAS meta-analysis data.

## Data Availability

All data produced in this work were simulated using data from the UK biobank. The access to the UK biobank data can be applied through https://www.ukbiobank.ac.uk/.
The scripts used to generate the data, along with an open source implementation of our method as an R package is available on GitHub.

https://github.com/JoonasKartau/finemapmiss

## 8 Acknowledgements

This research has been conducted using the UK Biobank Resource under Application Number 22627. We are grateful to Rashkin et al. and GWAS catalog for publicly available data and Pyörni Vartiainen for discussions.

