## Supplementary Material for "FINEMAP-miss: Fine-mapping genome-wide association studies with missing genotype information"

#### 1 Statistics of GWAS

**Notation:**

- $d$, number of datasets. A separate GWAS has been run on each dataset.
- $n$, number of individuals. $n_j$ the number of individuals at variant $j$. $n_{j\ell}$ the number of individuals with genotype information at both variants $j$ and $\ell$.
- $y$, quantitative phenotype value that has been standardized to have mean of 0 and standard deviation of 1.
- $p$, number of genetic variants in the region of interest.
- $g$, genotype value as an allele dosage of the effect allele. Can be either from genotyping (in $\{0, 1, 2\}$) or from imputation (in $[0, 2]$) or a missing value $\emptyset$
- $\boldsymbol{G}$, $n \times p$ matrix of genotype values with possibly missing genotype values.
- $\text{INFO}_j$, the information measure of variant $j$ after genotype imputation or summary statistics imputation (see definitions below).
- $I_j = \sqrt{\text{INFO}_j}$ and $\boldsymbol{I} = \text{diag}(I_j)$, the diagonal matrix of square roots of the INFO values.
- $\boldsymbol{X}$, $n \times p$ matrix of scaled genotype values, where each column of $\boldsymbol{G}$ has been mean-centered and scaled by standard deviation of the genotype in the population. Note: scaling is not done within the sample, where missingness and imperfect INFO ($< 1$) may exist, but rather it is done with respect to the reference population. Consequently, variance of column $j$ of $\boldsymbol{X}$ is $\text{INFO}_j$ and can be $< 1$. Assuming the Hardy-Weinberg equilibrium holds, the scaling for variant $j$ happens by dividing the mean-cenetered genotype dosage by $\sqrt{2f_j(1 - f_j)}$, where $f_j$ is the minor allele frequency at variant $j$.
- $x$, scaled genotype value per individual, or missing value. $x_j$ denotes a random variable of scaled genotype at variant $j$ while $x_{ij}$ is the value for individual $i$ at variant $j$, that is, $x_{ij} = \boldsymbol{X}_{ij}$, the element $(i,j)$ of matrix $\boldsymbol{X}$.
- $\boldsymbol{x}_j$, vector of scaled genotype values at variant $j$ over a set of individuals considered. The variance of the elements of $\boldsymbol{x}_j$ is $\text{INFO}_j$.
- $\boldsymbol{R}$, $p \times p$ linkage disequilibrium (LD) matrix, whose elements are observed correlation coefficients

$$\boldsymbol{R}_{j\ell} = \frac{\boldsymbol{x}_j{}^\intercal \boldsymbol{x}_\ell}{nI_jI_\ell}.$$

  Thus $\boldsymbol{R} = n^{-1}\boldsymbol{I}^{-1}\boldsymbol{X}^\intercal\boldsymbol{X}\boldsymbol{I}^{-1}$.

- $\boldsymbol{S}$, $p \times p$ diagonal matrix of GWAS standard errors $s_j$.
- $\boldsymbol{N} = \text{diag}(\sqrt{n_j})$, $p \times p$ diagonal matrix of the square root of the variant specific sample sizes.

**GWAS results for one study** contain variant-specific values

- $\widehat{\beta}_j$, marginal regression coefficient of standardized genotypes/dosages of variant $j$ from the univariate regression model $y = \mu + x_j\beta_j + \varepsilon$, namely,

$$\widehat{\beta}_j = \frac{\boldsymbol{x}_j{}^\intercal \boldsymbol{y}}{\boldsymbol{x}_j{}^\intercal \boldsymbol{x}_j}.$$

Note that when additional covariates, or more complex regression models, such as logistic regression or mixed effects models, are used, the marginal effect estimates from the GWAS software do not match exactly with the least squares formula given above. In that case, we will work with the actual GWAS output rather than the least squares solution.

- $s_j$, standard error (SE) of $\widehat{\beta}_j$, namely,

$$s_j = \frac{\widehat{\sigma}_\varepsilon}{\sqrt{\boldsymbol{x}_j{}^\intercal \boldsymbol{x}_j}},$$

where the residual variance is estimated as $\widehat{\sigma^2_\varepsilon} = 1 - \text{Var}(\boldsymbol{x}_j)\widehat{\beta}_j^2$, after $\text{Var}(y) = 1$. Note that if additional covariates or more complex regression models are used, SE may not match with the formula just given. In that case, we will work with the SE estimates from the actual GWAS output.

- $\widehat{z}_j = \widehat{\beta}_j / s_j$, is the z-score that can be computed from the effect estimates and their SEs.

Note that, typically, GWAS software produce the effect size estimates on the allelic scale, $\beta_j^{(g)}$, where genotype $g$ is the predictor in the regression model: $y = \mu + g_j\beta_j^{(g)} + \varepsilon$. Here, instead, we define the effects with respect to the scaled genotypes ($x$ is the predictor rather than $g$). One can transform between the two by the formula

$$\widehat{\beta}_j = \sqrt{\text{Var}(g_j)}\,\widehat{\beta}_j^{(g)} \approx \sqrt{2f_j(1-f_j)}\,\widehat{\beta}_j^{(g)},$$

where $f_j$ is the minor allele frequency of variant $j$ and the approximation is based on the assumption of the Hardy-Weinberg equilibrium. The same relationship holds for the SEs: $s_j \approx \sqrt{2f_j(1-f_j)}\,s_j^{(g)}$. The scaling is always done with respect to the expected population standard deviation of the genotype variable rather than with respect to the observed SD within the sample. The latter is typically smaller than the population SD when imputation INFO $< 1$.

Importantly, z-scores are not affected whether they were computed from the scaled effect sizes $\beta_j$ or from the allelic effect sizes $\beta_j^{(g)}$ since the constant of transformation is the same for both the effect sizes and for their SEs and thus cancels out from the z-scores.

#### 1.1 Genotype imputation

The genotype of a variant is not always measured by the genotyping array. Unobserved variants can be imputed using an imputation reference panel to obtain probabilistic estimates of the missing genotypes (e.g. using software series IMPUTE [1] or BEAGLE [2]). Essentially, imputation uses the dependencies (linkage disequilibrium (LD)) between the variants in the reference panel, together with the observed genotypes of the target sample in neighboring variants, to predict the genotypes at the missing variants in the target sample. For individual $i$ and variant $j$, imputation produces a set of 3 probabilities pertaining to the 3 possible genotypes:

$$p\left(G_{ij} = g\right) = \begin{cases} p_{ij,0} & g = 0, \\ p_{ij,1} & g = 1, \\ p_{ij,2} & g = 2. \end{cases}$$

With the genotype probabilities, it is possible to construct a genotype dosage $\overline{g_{ij}}$ as the expected genotype given the imputation distribution:

$$\overline{g_{ij}} = \mathbb{E}[G_{ij}|\boldsymbol{p}_{ij}] = 0 \cdot p_{ij,0} + 1 \cdot p_{ij,1} + 2 \cdot p_{ij,2},$$

which is substituted as the genotype when performing a GWAS.

For a fixed variant, $j$, let's consider the variance of the genotype across the individuals, considered as a random variable for which the imputation distribution $\boldsymbol{p}_{ij}$ is available for all individuals $i$. The law of total variance gives

$$\mathrm{Var}(G_{ij}) = \mathbb{E}[\mathrm{Var}(G_{ij}|\boldsymbol{p}_{ij})] + \mathrm{Var}[\mathbb{E}(G_{ij}|\boldsymbol{p}_{ij})] = \mathbb{E}[\mathrm{Var}(G_{ij}|\boldsymbol{p}_{ij})] + \mathrm{Var}(\overline{g_{ij}}).$$

$\mathbb{E}[\mathrm{Var}(G_{ij}|\boldsymbol{p}_{ij})] = \mathrm{Var}(G_{ij}) - \mathrm{Var}(\overline{g_{ij}})$ describes the average uncertainty caused by the imputation compared to the case where every genotype were observed exactly, in which case this quantity would be 0. Thus, the closer $\mathrm{Var}(\overline{g_{ij}})$ is to $\mathrm{Var}(G_{ij})$ the more informative the imputation at the variant $j$ is. Imputation INFO (and its square root $I$) is defined as

$$\mathrm{INFO}_j = I_j^2 = \frac{\mathrm{Var}(\overline{g_{ij}})}{\mathrm{Var}(G_{ij})} = 1 - \frac{\mathbb{E}[\mathrm{Var}(G_{ij}|\boldsymbol{p}_{ij})]}{\mathrm{Var}(G_{ij})} = 1 - \frac{\sum_{i=1}^{n_j} v_{ij}}{n_j V_j},$$

where $V_j$ is the variance of the genotype at variant $j$ in the population and $v_{ij}$ is the variance of the imputation distribution of individual $i$ at variant $j$:

$$\begin{aligned} v_{ij} &= \mathrm{Var}[G_{ij}|\boldsymbol{p}_{ij}] = \mathbb{E}[G_{ij}^2|\boldsymbol{p}_{ij}] - \mathbb{E}[G_{ij}|\boldsymbol{p}_{ij}]^2, \\ &= 4 \cdot p_{ij,2} + 1 \cdot p_{ij,1} + 0 \cdot p_{ij,0} - (2 \cdot p_{ij,2} + 1 \cdot p_{ij,1} + 0 \cdot p_{ij,0})^2. \end{aligned}$$

Assuming the Hardy-Weinberg equilibrium, $V_j \approx 2f_j(1 - f_j)$, where $f_j$ is the MAF at variant $j$. When INFO is 1, there is no uncertainty due to imputation and when INFO is 0, the imputation is completely uninformative as it proposes the same genotype dosage for every individual, independent of the observed genotypes at other variants.

As a result of genotype imputation, the variance of the dosage data is shrunk by a factor INFO compared to the variance of the true genotypes:

$$\mathrm{Var}\left(\overline{g_{ij}}\right) = I_j^2 \cdot \mathrm{Var}\left(G_{ij}\right).$$

##### Effect sizes with imputation

As explained above, we consider the GWAS effects $\beta_j$ estimated with respect to the scaled genotype $x$, i.e., the mean centered genotype value scaled by the standard deviation of the genotype in the population, rather than with respect to $g$, the genotype variable on the allelic scale (0,1,2). Note that the scaling of the genotypes is done with respect to the standard deviation in the population while INFO is not affecting the scaling. Thus, at variant $j$, the variance of the scaled genotype $x_{\cdot j}$ over the individuals is $\mathrm{INFO}_j$, which is $< 1$ if there is uncertainty related to imputation at the variant.

Consider one variant. Suppose that it were possible to compare the GWAS summary statistics $(\beta, s)$ from regression on imputed (scaled) dosage data $x$ to results $(\beta^*, s^*)$ from regression on fully observed (scaled) genotype data $x^*$. We assume that the imputation distributions $\boldsymbol{p}_i = (p_{i,0}, p_{i_1}, p_{i,2})$ for all individuals $i$ are correctly calibrated, i.e., the conditional distribution $x^*|\boldsymbol{p}_i$ correctly describes the true genotype distribution of individual $i$.

By definition, the dosage variable is $x_i = \mathbb{E}(x^*|\boldsymbol{p}_i)$. By the tower property of conditional expectation the expected scaled genotype in the population is $\mathbb{E}(x^*) = \mathbb{E}(\mathbb{E}(x^*|\boldsymbol{p}_i)) = \mathbb{E}(x)$, where the expectation is over the individuals for whom we have imputation information. To find the covariance between $x$ and $x^*$, note that

$$\begin{aligned} \mathbb{E}(xx^*) &= \mathbb{E}(\mathbb{E}(xx^*|\boldsymbol{p}_i)) \\ &= \mathbb{E}(x\mathbb{E}(x^*|\boldsymbol{p}_i)) \\ &= \mathbb{E}(x^2). \end{aligned}$$

Hence, the covariance is

$$\mathrm{Cov}(x, x^*) = \mathbb{E}(xx^*) - \mathbb{E}(x)\mathbb{E}(x^*) = \mathbb{E}(x^2) - \mathbb{E}(x)^2 = \mathrm{Var}(x) = \mathrm{INFO}$$

and correlation $\mathrm{Cor}(x, x^*) = \sqrt{\mathrm{INFO}} = I$.

To study the effect sizes from linear regression, write the phenotype $y = \mu + x^*\beta^* + \varepsilon$, where the effect size for the fully observed scaled genotype $x^*$ is $\beta^*$. When we regress $y$ on $x$, the regression coefficient is

$$\beta = \frac{\mathrm{Cov}(y,x)}{\mathrm{Var}(x)} = \frac{\mathrm{Cov}(x^*\beta^* + \varepsilon, x)}{\mathrm{Var}(x)} = \beta^* \frac{\mathrm{Cov}(x^*, x)}{\mathrm{Var}(x)} = \beta^* \frac{\mathrm{Var}(x)}{\mathrm{Var}(x)} = \beta^*.$$

Hence, we expect that a correctly calibrated genotype imputation does not cause bias in the effect size estimate of the variant. However, it does inflate the standard error, because the variance of the imputed dosage is smaller than the variance of the genotype variable whenever INFO $< 1$.

In conclusion, we expect the following relationships for variant $j$:

$$\begin{aligned}
\mathbb{E}\left[\widehat{\beta}_j\right] &= \mathbb{E}\left[\widehat{\beta}_j^*\right] \\
s_j &\approx \sqrt{\frac{1 - I_j^2\left(\widehat{\beta}_j\right)^2}{n_j I_j^2}} \\
s_j^* &\approx \sqrt{\frac{1 - \left(\widehat{\beta}_j^*\right)^2}{n_j}}.
\end{aligned}$$

#### 1.2 Covariance of effect estimators within one study

Consider two variants 1 and 2, where GWAS results have been computed using regression on $n_1$ and $n_2$ individuals, respectively, with $n_{12}$ overlapping individuals. Denote by $r_{12}$ the correlation of the scaled genotype dosages $x_1$ and $x_2$ in the population. Here, the scaling is by the population standard deviation, approximately $\sqrt{2f_j(1-f_j)}$, where $f_j$ is the minor allele frequency of variant $j$. Note that this scaling does not result in each $x_j$ being standardized to have variance of 1, but rather its variance equals to $I_j^2 \leq 1$.

Given the true effect sizes $\beta_1$ and $\beta_2$ in the population, we can explain the phenotype $y$ by two separate univariate regression models depending on which variant we consider

$$y = x_1\beta_1 + \varepsilon_1 \text{ or } y = x_2\beta_2 + \varepsilon_2.$$

Below we treat the scaled dosages $x_1$ and $x_2$ and the error terms $\varepsilon_1$ and $\varepsilon_2$ as random variables. We use the bracket notation as a shorthand for covariance operator, thus, $[u, v] = \mathrm{Cov}(u, v)$ for any random variables $u$ and $v$. We have following properties:

- $[x_1, \varepsilon_1] = [x_2, \varepsilon_2] = 0$ because we assume that the linear model holds and that the predictors are uncorrelated with the error terms. Indeed, in the fitted linear model, the corresponding property always holds for the observed predictors and residuals.

- The two error terms are correlated with covariance

$$\begin{aligned}
[\varepsilon_1, \varepsilon_2] &= [y - x_1\beta_1, y - x_2\beta_2] \\
&= [y, y] - \beta_1[x_1, y] - \beta_2[y, x_2] + \beta_1\beta_2[x_1, x_2] \\
&= 1 - \beta_1^2 I_1^2 - \beta_2^2 I_2^2 + \beta_1\beta_2 r_{12} I_1 I_2 \qquad (1)
\end{aligned}$$

- $\boldsymbol{x}_j^\intercal \boldsymbol{x}_j = \sum_{i=1}^{n_j} x_{ij}^2 \approx n_j \mathrm{Var}(x_j) = n_j I_j^2$ for both variants $j = 1, 2$.

Let us write the covariance between the effect size estimators of variants 1 and 2. Denote by $S_1$ and $S_2$ the sets of individuals that have data on variant 1 and 2, respectively. Then $|S_1| = n_1$, $|S_2| = n_2$ and $|S_1 \cap S_2| = n_{12}$.

$$
\begin{aligned}
\left[\widehat{\beta}_1, \widehat{\beta}_2\right] &= \left[\frac{\boldsymbol{x}_1^\intercal \boldsymbol{y}}{\boldsymbol{x}_1^\intercal \boldsymbol{x}_1}, \frac{\boldsymbol{x}_2^\intercal \boldsymbol{y}}{\boldsymbol{x}_2^\intercal \boldsymbol{x}_2}\right] \\
&= \left[\frac{\boldsymbol{x}_1^\intercal (\boldsymbol{x}_1 \beta_1 + \boldsymbol{\varepsilon}_1)}{\boldsymbol{x}_1^\intercal \boldsymbol{x}_1}, \frac{\boldsymbol{x}_2^\intercal (\boldsymbol{x}_2 \beta_2 + \boldsymbol{\varepsilon}_2)}{\boldsymbol{x}_2^\intercal \boldsymbol{x}_2}\right] \\
&= \left[\beta_1 + \frac{\boldsymbol{x}_1^\intercal \boldsymbol{\varepsilon}_1}{n_1 I_1^2}, \beta_2 + \frac{\boldsymbol{x}_2^\intercal \boldsymbol{\varepsilon}_2}{n_2 I_2^2}\right] \\
&= \frac{1}{n_1 n_2 I_1^2 I_2^2} \left[\boldsymbol{x}_1^\intercal \boldsymbol{\varepsilon}_1, \boldsymbol{x}_2^\intercal \boldsymbol{\varepsilon}_2\right] \\
&= \frac{1}{n_1 n_2 I_1^2 I_2^2} \left[\sum_{i \in S_1} x_{i1} \varepsilon_{i1}, \sum_{i \in S_2} x_{i2} \varepsilon_{i2}\right] \\
&= \frac{1}{n_1 n_2 I_1^2 I_2^2} \sum_{i \in S_1 \cap S_2} \left[x_{i1} \varepsilon_{i1}, x_{i2} \varepsilon_{i2}\right] \\
&\approx \frac{n_{12}}{n_1 n_2 I_1^2 I_2^2} \left[x_1 \varepsilon_1, x_2 \varepsilon_2\right] \\
&= \frac{n_{12}}{n_1 n_2 I_1^2 I_2^2} \left(\mathbb{E}(x_1 \varepsilon_1 x_2 \varepsilon_2) - \mathbb{E}(x_1 \varepsilon_1) \mathbb{E}(x_2 \varepsilon_2)\right) && (2) \\
&= \frac{n_{12}}{n_1 n_2 I_1^2 I_2^2} \left(\mathbb{E}\left(\mathbb{E}(x_1 \varepsilon_1 x_2 \varepsilon_2 \,|\, x_1, x_2)\right) - 0 \cdot 0\right) && (3) \\
&= \frac{n_{12}}{n_1 n_2 I_1^2 I_2^2} \mathbb{E}\left(x_1 x_2 \mathbb{E}(\varepsilon_1 \varepsilon_2)\right) \\
&= \frac{n_{12}}{n_1 n_2 I_1^2 I_2^2} \mathbb{E}\left(x_1 x_2 \left(1 - \beta_1^2 I_1^2 - \beta_2^2 I_2^2 + \beta_1 \beta_2 r_{12} I_1 I_2\right)\right) && (4) \\
&= \frac{n_{12}}{n_1 n_2 I_1^2 I_2^2} r_{12} I_1 I_2 \left(1 - \beta_1^2 I_1^2 - \beta_2^2 I_2^2 + \beta_1 \beta_2 r_{12} I_1 I_2\right) && (5) \\
&= \frac{n_{12}}{n_1 n_2 I_1 I_2} r_{12} \left(1 - \beta_1^2 I_1^2 - \beta_2^2 I_2^2 + \beta_1 \beta_2 r_{12} I_1 I_2\right) && (6)
\end{aligned}
$$

Above, step (2) is the definition of covariance; (3) is the law of total expectation and the uncorrelatedness between $x_j$ and $\varepsilon_j$; (4) is the covariance of error terms from equation (1) and (5) is the fact that $\mathbb{E}(x_1 x_2) = [x_1, x_2] = r_{12} I_1 I_2$. For z-scores, we have

$$
[\widehat{z}_1, \widehat{z}_2] = \left[\frac{\widehat{\beta}_1}{s_1}, \frac{\widehat{\beta}_2}{s_2}\right] = \frac{n_{12}}{n_1\, n_2 I_1 I_2\, s_1\, s_2} r_{12} \left(1 - \beta_1^2 I_1^2 - \beta_2^2 I_2^2 + \beta_1 \beta_2 r_{12} I_1 I_2\right). \tag{7}
$$

Theoretically, each standard error above is given as

$$
s_j = \frac{\sigma_{\varepsilon_j}}{\sqrt{n_j I_j^2}} = \sqrt{\frac{1 - I_j^2 \beta_j^2}{n_j I_j^2}},
$$

so that

$$
[\widehat{z}_1, \widehat{z}_2] = \frac{n_{12}}{\sqrt{n_1 n_2} \sqrt{1 - \beta_1^2 I_1^2} \sqrt{1 - \beta_2^2 I_1^2}} r_{12} \left(1 - \beta_1^2 I_1^2 - \beta_2^2 I_1^2 + \beta_1 \beta_2 r_{12} I_1 I_2\right). \tag{8}
$$

In practice, we may use the empirical SE estimates given by the GWAS software to account for deviations from the theoretical model.

Note that the covariance between the z-scores is also the correlation between the marginal effects, $\mathrm{Cor}\left(\widehat{\beta}_1, \widehat{\beta}_2\right)$.

If we consider estimating the effects of $k$ variants jointly using a multiple regression model instead of the univariate marginal model that was used above, then all variants must have been observed on the same set of $n$ individuals. Our multiple regression model is

$$
y = \mu + x_1 \lambda_1 + \ldots + x_k \lambda_k + \varepsilon_\lambda,
$$

where we use $\lambda$ to denote the effect from the joint model instead of $\beta$ that was used for the marginal effect. The theory of multiple linear regression tells us that

$$\widehat{\boldsymbol{\lambda}} = \left(\boldsymbol{X}^\intercal \boldsymbol{X}\right)^{-1} \boldsymbol{X}^\intercal \boldsymbol{y}.$$

To write the estimates of the joint effects $\widehat{\boldsymbol{\lambda}}$ as a function of the marginal effects $\widehat{\boldsymbol{\beta}} = (n\boldsymbol{I}^2)^{-1}\boldsymbol{X}^\intercal\boldsymbol{y}$ from GWAS data, we have

$$\begin{aligned}
\widehat{\boldsymbol{\lambda}} &= \left(\boldsymbol{X}^\intercal \boldsymbol{X}\right)^{-1} \boldsymbol{X}^\intercal \boldsymbol{y} \\
&= \boldsymbol{I}^{-1} \left(\boldsymbol{I}^{-1}\boldsymbol{X}^\intercal \boldsymbol{X}\boldsymbol{I}^{-1}\right)^{-1} \boldsymbol{I}^{-1}\boldsymbol{X}^\intercal \boldsymbol{y} \\
&= n\boldsymbol{I}^{-1} \left(\boldsymbol{I}^{-1}\boldsymbol{X}^\intercal \boldsymbol{X}\boldsymbol{I}^{-1}\right)^{-1} \boldsymbol{I}^{-1}\frac{\boldsymbol{X}^\intercal \boldsymbol{y}}{n} \\
&= \boldsymbol{I}^{-1}\boldsymbol{R}^{-1}\boldsymbol{I}^{-1}\frac{\boldsymbol{X}^\intercal \boldsymbol{y}}{n} \\
&= \boldsymbol{I}^{-1}\boldsymbol{R}^{-1}\boldsymbol{I}\boldsymbol{I}^{-1}\boldsymbol{I}^{-1}\frac{\boldsymbol{X}^\intercal \boldsymbol{y}}{n} \\
&= \boldsymbol{I}^{-1}\boldsymbol{R}^{-1}\boldsymbol{I}\widehat{\boldsymbol{\beta}}
\end{aligned}$$

where $\widehat{\boldsymbol{\beta}}$ contains the marginal effect estimates of the $k$ variants and $\boldsymbol{R} = n^{-1}\boldsymbol{I}^{-1}\boldsymbol{X}^\intercal\boldsymbol{X}\boldsymbol{I}^{-1}$ is their $k \times k$ matrix of correlations of genotypes / genotype dosages called the LD matrix. Note that even though imperfect ($I_j < 1$) but correctly calibrated imputation does not cause bias in the expected marginal effects $\widehat{\boldsymbol{\beta}}$, it does modify the expected joint effects, which under the complete information would equal to $\boldsymbol{\lambda} = \boldsymbol{R}^{-1}\boldsymbol{\beta}$. We can estimate the residual variance using the estimates of the marginal effects as

$$\sigma^2_{\varepsilon_\lambda} = 1 - \mathrm{Var}(\boldsymbol{X}\boldsymbol{\lambda}) \approx 1 - n^{-1}\widehat{\boldsymbol{\lambda}}^\intercal\boldsymbol{X}^\intercal\boldsymbol{X}\widehat{\boldsymbol{\lambda}} = 1 - \widehat{\boldsymbol{\lambda}}^\intercal\boldsymbol{I}\boldsymbol{R}\boldsymbol{I}\widehat{\boldsymbol{\lambda}} \approx 1 - \widehat{\boldsymbol{\beta}}^\intercal\boldsymbol{I}\boldsymbol{R}^{-1}\boldsymbol{I}\widehat{\boldsymbol{\beta}}.$$

The covariance matrix of the estimator of joint effects is

$$\mathrm{Cov}\left(\widehat{\boldsymbol{\lambda}}\right) = \left(\boldsymbol{X}^\intercal \boldsymbol{X}\right)^{-1}\sigma^2_{\varepsilon_\lambda} \approx \boldsymbol{I}^{-1}\left(n\boldsymbol{R}\right)^{-1}\boldsymbol{I}^{-1}(1 - \widehat{\boldsymbol{\lambda}}^\intercal\boldsymbol{I}\boldsymbol{R}\boldsymbol{I}\widehat{\boldsymbol{\lambda}}) \approx \frac{(1 - \widehat{\boldsymbol{\beta}}^\intercal\boldsymbol{I}\boldsymbol{R}^{-1}\boldsymbol{I}\widehat{\boldsymbol{\beta}})}{n}\boldsymbol{I}^{-1}\boldsymbol{R}^{-1}\boldsymbol{I}^{-1}.$$

#### 1.3 Covariance of effect estimators in meta-analysis

Suppose the GWAS effect estimates result from a fixed-effects meta-analysis across $d$ independent GWAS, where the meta-analysis method is inverse variance weighting (IVW). Let $\widehat{\beta}_{ju}$ be the marginal effect estimate of the $j$th variant in the $u$th study, and $s_{ju}$ be the corresponding standard error. IVW meta-analysis uses the precision (defined as the inverse of the variance) of the effect estimate as the weight $w_{ju} = s_{ju}^{-2}$ to combine the data into the meta-analyzed marginal effect $\widehat{\beta}_j^{(m)}$, standard error $s_j^{(m)}$, and z-score $\widehat{z}_j^{(m)}$:

$$\begin{aligned}
\widehat{\beta}_j^{(m)} &= \frac{w_{j1}\widehat{\beta}_{j1} + \cdots + w_{jd}\widehat{\beta}_{jd}}{w_{j1} + \cdots + w_{jd}} \\
s_j^{(m)} &= (w_{j1} + \cdots + w_{jd})^{-\frac{1}{2}} \\
\widehat{z}_j^{(m)} = \frac{\widehat{\beta}_j^{(m)}}{s_j^{(m)}} &= \frac{w_{j1}\widehat{\beta}_{j1} + \cdots + w_{jd}\widehat{\beta}_{jd}}{\sqrt{w_{j1} + \cdots + w_{jd}}}.
\end{aligned}$$

To account for missing data, we agree that if variant $j$ is not observed in study $u$, then we set $w_{ju} = 0$. Implicitly we assume that each variant is observed in at least one study.

Consider the covariance of meta-analyzed z-scores at two variants $j_1$ and $j_2$,

$$\begin{aligned}
\mathrm{Cov}\left(\widehat{z}_{j_1}^{(m)}, \widehat{z}_{j_2}^{(m)}\right) &= \mathrm{Cov}\left(\frac{\sum_{u=1}^{d} w_{j_1 u}\widehat{\beta}_{j_1 u}}{\sqrt{\sum_{u=1}^{d} w_{j_1 u}}}, \frac{\sum_{v=1}^{d} w_{j_2 v}\widehat{\beta}_{j_2 v}}{\sqrt{\sum_{v=1}^{d} w_{j_2 v}}}\right) \\
&= \frac{\sum_{u=1}^{d}\sum_{v=1}^{d} w_{j_1 u} w_{j_2 v}\mathrm{Cov}\left(\widehat{\beta}_{j_1 u}, \widehat{\beta}_{j_2 v}\right)}{\sqrt{\sum_{u=1}^{d} w_{j_1 u}}\sqrt{\sum_{v=1}^{d} w_{j_2 v}}} \\
&= \frac{\sum_{u=1}^{d} w_{j_1 u} w_{j_2 u}\mathrm{Cov}\left(\widehat{\beta}_{j_1 u}, \widehat{\beta}_{j_2 u}\right)}{\sqrt{\sum_{u=1}^{d} w_{j_1 u}}\sqrt{\sum_{u=1}^{d} w_{j_2 u}}}.
\end{aligned}$$

The double sum is lost on row 3, since datasets are assumed to be independent, that is, there are no overlapping or closely related individuals between any pairs of the studies. We can now use formula (6) to estimate $\mathrm{Cov}\left(\widehat{\beta}_{j_1 u}, \widehat{\beta}_{j_2 u}\right)$. If we assume that at each component study $u$, both variants $j_1$ and $j_2$ were observed (possibly after imputation) on the full sample of size $n_u$, then we have an approximation

$$\begin{aligned}
\mathrm{Cov}\left(\widehat{\beta}_{j_1 u}, \widehat{\beta}_{j_2 u}\right) &\approx \frac{\left(1 - \widehat{\beta}_{j_1 u}^2 I_{j_1 u}^2 - \widehat{\beta}_{j_2 u}^2 I_{j_2 u}^2 + \widehat{\beta}_{j_1 u}\widehat{\beta}_{j_2 u} r_{j_1 j_2, u} I_{j_1 u} I_{j_2 u}\right)}{n_u I_{j_1 u} I_{j_2 u}} r_{j_1 j_2, u} \\
&= \widehat{s}_{j_1 u}\widehat{s}_{j_2 u}\frac{\left(1 - \widehat{\beta}_{j_1 u}^2 I_{j_1 u}^2 - \widehat{\beta}_{j_2 u}^2 I_{j_2 u}^2 + \widehat{\beta}_{j_1 u}\widehat{\beta}_{j_2 u} r_{j_1 j_2, u} I_{j_1 u} I_{j_2 u}\right)}{\sqrt{1 - I_{j_1 u}^2 \widehat{\beta}_{j_1 u}^2}\sqrt{1 - I_{j_2 u}^2 \widehat{\beta}_{j_2 u}^2}} r_{j_1 j_2, u} \\
&= \frac{\left(1 - \widehat{\beta}_{j_1 u}^2 I_{j_1 u}^2 - \widehat{\beta}_{j_2 u}^2 I_{j_2 u}^2 + \widehat{\beta}_{j_1 u}\widehat{\beta}_{j_2 u} r_{j_1 j_2, u} I_{j_1 u} I_{j_2 u}\right)}{\sqrt{w_{j_1 u} w_{j_2 u}}\sqrt{1 - I_{j_1 u}^2 \widehat{\beta}_{j_1 u}^2}\sqrt{1 - I_{j_2 u}^2 \widehat{\beta}_{j_2 u}^2}} r_{j_1 j_2, u}, \\
&= \frac{B_{j_1 j_2, u}}{\sqrt{w_{j_1 u} w_{j_2 u}}} r_{j_1 j_2, u},
\end{aligned}$$

where $r_{j_1 j_2, u}$ is the correlation between the scaled genotypes/dosages of variants $j_1$ and $j_2$ in study $u$, and we have defined

$$B_{j_1 j_2, u} = \frac{\left(1 - \widehat{\beta}_{j_1 u}^2 I_{j_1 u}^2 - \widehat{\beta}_{j_2 u}^2 I_{j_2 u}^2 + \widehat{\beta}_{j_1 u}\widehat{\beta}_{j_2 u} r_{j_1 j_2, u} I_{j_1 u} I_{j_2 u}\right)}{\sqrt{1 - I_{j_1 u}^2 \widehat{\beta}_{j_1 u}^2}\sqrt{1 - I_{j_2 u}^2 \widehat{\beta}_{j_2 u}^2}}.$$

On the row 2 above, we prefer to use the observed SEs ($\widehat{s}_{j_1 u}$ and $\widehat{s}_{j_2 u}$) rather than the theoretical effective sample size factor $n_u I_{j_1 u} I_{j_2 u}$ in the computations to account for possible discrepancies between theoretical and effective sample sizes, which also results in additional stability when the covariance matrix is inverted. With these assumptions, we have that the correlation $r_{j_1 j_2}^{(m)}$ of the meta-analyzed z-scores is

$$\begin{aligned}
r_{j_1 j_2}^{(m)} &= \mathrm{Cor}\left(\widehat{z}_{j_1}^{(m)}, \widehat{z}_{j_2}^{(m)}\right) = \mathrm{Cov}\left(\widehat{z}_{j_1}^{(m)}, \widehat{z}_{j_2}^{(m)}\right) \\
&= \frac{\sum_{u=1}^{d} w_{j_1 u} w_{j_2 u}\frac{B_{j_1 j_2, u}}{\sqrt{w_{j_1 u} w_{j_2 u}}} r_{j_1 j_2, u}}{\sqrt{\sum_{u=1}^{d} w_{j_1 u}}\sqrt{\sum_{u=1}^{d} w_{j_2 u}}} \approx r_{j_1 j_2} \cdot \frac{\sum_{u=1}^{d}\sqrt{w_{j_1 u} w_{j_2 u}} B_{j_1 j_2, u}}{\sqrt{\sum_{u=1}^{d} w_{j_1 u}}\sqrt{\sum_{u=1}^{d} w_{j_2 u}}},
\end{aligned}$$

where the approximation on the last line is applicable when the correlation $r_{j_1 j_2}$ between the genotypes/dosages is similar across the studies. We denote the the covariance matrix of the meta-analyzed z-scores as

$$\boldsymbol{R_M} = \left(r_{j_1 j_2}^{(m)}\right).$$

We emphasize that $\boldsymbol{R_M}$ is different from the LD matrix $\boldsymbol{R} = (r_{j_1 j_2})$.

##### 1.4 Relationship between joint effects and meta-analyzed marginal effects.

Define $\boldsymbol{\lambda}(\boldsymbol{\beta}) := \boldsymbol{I}^{-1}\boldsymbol{R}^{-1}\boldsymbol{I}\boldsymbol{\beta}$ to be the function that maps the marginal effects $\boldsymbol{\beta}$ to the joint ("causal") effects $\boldsymbol{\lambda}$, as derived above in section 1.2. We assume that if the likelihood function for a given meta-analyzed data set is proportional to a Gaussian density centered at $\widehat{\boldsymbol{\beta}}^{(m)}$, then the likelihood function for the joint effects is achieved by applying the linear transformation $\boldsymbol{\lambda}(\boldsymbol{\beta})$, i.e., the maximum likelihood value is

$$\widehat{\boldsymbol{\lambda}} = \boldsymbol{\lambda}(\widehat{\boldsymbol{\beta}}^{(m)}) = \boldsymbol{I}^{-1}\boldsymbol{R}^{-1}\boldsymbol{I}\widehat{\boldsymbol{\beta}}^{(m)} = \boldsymbol{I}^{-1}\boldsymbol{R}^{-1}\boldsymbol{I}\boldsymbol{S}\widehat{\boldsymbol{z}}^{(m)}, \tag{9}$$

where $\boldsymbol{S}$ is the $p \times p$ diagonal matrix of GWAS standard errors $\widehat{s}_j$. We will denote the variance of the transformed MLE by $\boldsymbol{\Theta}$,

$$\begin{aligned}\boldsymbol{\Theta} := \mathrm{Var}(\widehat{\boldsymbol{\lambda}}) &= \boldsymbol{I}^{-1}\boldsymbol{R}^{-1}\boldsymbol{I}\boldsymbol{S}\,\mathrm{Var}(\widehat{\boldsymbol{z}}^{(m)})\,\boldsymbol{S}\boldsymbol{I}\boldsymbol{R}^{-1}\boldsymbol{I}^{-1}, \\ &= \boldsymbol{I}^{-1}\boldsymbol{R}^{-1}\boldsymbol{I}\boldsymbol{S}\boldsymbol{R}_{\boldsymbol{M}}\boldsymbol{S}\boldsymbol{I}\boldsymbol{R}^{-1}\boldsymbol{I}^{-1}. \end{aligned} \tag{10}$$

#### 2 Deriving the marginal likelihood of causal configurations in meta-analyses

The derivations below are applicable for GWAS summary statistics from meta-analyses and, for notational simplicity, we omit the specific subscript $(m)$ that previously denoted that the estimates were derived from a meta-analysis.

##### 2.1 Likelihood and prior for joint effects

As explained in the main text, we use an extension of the FINEMAP model of Benner et al. [3] to define the model for causal configurations. A causal configuration $\boldsymbol{\gamma} \in \{0,1\}^p$ is a (sparse) binary vector indicating which of the $p$ genetic variants are considered causal, i.e., having non-zero effects in the joint model. Given the data DATA, that could be either the original individual-level genotype-phenotype data or GWAS summary statistics, it is possible to evaluate the support that DATA give for a configuration $\boldsymbol{\gamma}$ using the marginal likelihood $L(\boldsymbol{\gamma}; \mathrm{DATA}) = p(\mathrm{DATA}|\boldsymbol{\gamma})$. In the Bayesian framework, the marginal likelihood is

$$L(\boldsymbol{\gamma}; \mathrm{DATA}) = p(\mathrm{DATA}|\boldsymbol{\gamma}) = \int p(\mathrm{DATA}|\boldsymbol{\lambda})\, p(\boldsymbol{\lambda}|\boldsymbol{\gamma})\, \mathrm{d}\boldsymbol{\gamma},$$

where $p(\mathrm{DATA}|\boldsymbol{\lambda})$ is the likelihood function of the causal effects and $p(\boldsymbol{\lambda}|\boldsymbol{\gamma})$ is the prior probability distribution of the effects $\boldsymbol{\lambda}$ under the causal configuration $\boldsymbol{\gamma}$.

For a given configuration $\boldsymbol{\gamma}$, let $\Gamma_{\boldsymbol{\gamma}} \subset \{1,\ldots,p\}$ be the set of its causal variants and $k_{\boldsymbol{\gamma}} = |\Gamma_{\boldsymbol{\gamma}}|$. We assume a priori that if a variant $j$ is causal ($j \in \Gamma_{\boldsymbol{\gamma}}$), then its effect is an independent, Gaussian random variable with mean 0 and variance $\tau^2$, and if $j$ is non-causal ($j \notin \Gamma_{\boldsymbol{\gamma}}$), then its effect is 0. Here, $\tau^2 > 0$ is given as an input parameter to the model. Setting the effects of the non-causal variants to 0 results in a degenerate prior probability distribution. To rectify this during the theoretical derivations, we set a non-zero variance $v > 0$ for the non-causal effects, and take the limit $v \to 0$ after we have simplified the formulas. The multivariate prior distribution for $\boldsymbol{\lambda}|\boldsymbol{\gamma}$ is thus defined by the diagonal covariance matrix $\boldsymbol{\Sigma}_{\boldsymbol{\gamma}}$ whose element $j$ on the diagonal is

$$\mathrm{diag}(\boldsymbol{\Sigma}_{\boldsymbol{\gamma}})_j = \begin{cases} \tau^2, & \text{if } j \in \Gamma_{\boldsymbol{\gamma}} \\ v, & \text{if } j \notin \Gamma_{\boldsymbol{\gamma}}. \end{cases}$$

By combining the prior $p(\boldsymbol{\lambda}|\boldsymbol{\gamma}) = \mathcal{N}(\boldsymbol{\lambda}|\boldsymbol{0}_p, \boldsymbol{\Sigma}_{\boldsymbol{\gamma}})$ with the likelihood $L(\boldsymbol{\lambda}; \mathrm{DATA}) \propto \mathcal{N}(\boldsymbol{\lambda}|\widehat{\boldsymbol{\lambda}}, \boldsymbol{\Theta})$, we have

$$
\begin{aligned}
L(\boldsymbol{\gamma}; \text{DATA}) &= \int p(\text{DATA}|\boldsymbol{\lambda}, \boldsymbol{\gamma}) \cdot p(\boldsymbol{\lambda}|\boldsymbol{\gamma}) d\boldsymbol{\lambda}, \\
&\propto \int \mathcal{N}(\boldsymbol{\lambda}|\widehat{\boldsymbol{\lambda}}, \boldsymbol{\Theta}) \cdot \mathcal{N}(\boldsymbol{\lambda}|\mathbf{0}_p, \boldsymbol{\Sigma}_{\boldsymbol{\gamma}}) d\boldsymbol{\lambda}, \\
&= \int \mathcal{N}(\widehat{\boldsymbol{\lambda}}|\boldsymbol{\lambda}, \boldsymbol{\Theta}) \cdot \mathcal{N}(\boldsymbol{\lambda}|\mathbf{0}_p, \boldsymbol{\Sigma}_{\boldsymbol{\gamma}}) d\boldsymbol{\lambda}, \\
&= \mathcal{N}(\widehat{\boldsymbol{\lambda}}|\mathbf{0}_p, \boldsymbol{\Theta} + \boldsymbol{\Sigma}_{\boldsymbol{\gamma}}) \\
&= \mathcal{N}(\widehat{\boldsymbol{\lambda}}|\mathbf{0}_p, \boldsymbol{I}^{-1}\boldsymbol{R}^{-1}\boldsymbol{I}\boldsymbol{S}\boldsymbol{R}_{\boldsymbol{M}}\boldsymbol{S}\boldsymbol{I}\boldsymbol{R}^{-1}\boldsymbol{I}^{-1} + \boldsymbol{\Sigma}_{\boldsymbol{\gamma}}) \\
&= \mathcal{N}(\widehat{\boldsymbol{z}}|\mathbf{0}_p, \boldsymbol{R}_{\boldsymbol{M}} + \boldsymbol{S}^{-1}\boldsymbol{I}^{-1}\boldsymbol{R}\boldsymbol{I}\boldsymbol{\Sigma}_{\boldsymbol{\gamma}}\boldsymbol{I}\boldsymbol{R}\boldsymbol{I}^{-1}\boldsymbol{S}^{-1}).
\end{aligned}
$$

Here the last line follows from the transformation (9). Thus, the marginal likelihood of $\boldsymbol{\gamma}$ in a meta-analysis is proportional to a multivariate normal density function evaluated at the observed z-scores. We can now drop the dimension by taking the limit as the non-causal prior variance $v \to 0$:

$$
L(\boldsymbol{\gamma}; \text{DATA}) \propto \lim_{v \to 0} \mathcal{N}(\widehat{\boldsymbol{z}}|\mathbf{0}_p, \boldsymbol{R}_{\boldsymbol{M}} + \boldsymbol{S}^{-1}\boldsymbol{I}^{-1}\boldsymbol{R}\boldsymbol{I}\boldsymbol{\Sigma}_{\boldsymbol{\gamma}}\boldsymbol{I}\boldsymbol{R}\boldsymbol{I}^{-1}\boldsymbol{S}^{-1}).
$$

As we take the limit, only the diagonal indices $j \in \Gamma_{\boldsymbol{\gamma}}$ remain non-zero in $\boldsymbol{\Sigma}_{\gamma}$ and any index $j' \notin \Gamma_{\boldsymbol{\gamma}}$ can be ignored, simplifying the second part of the covariance matrix to the form

$$
\lim_{v \to 0} \boldsymbol{S}^{-1}\boldsymbol{I}^{-1}\boldsymbol{R}\boldsymbol{I}\boldsymbol{\Sigma}_{\boldsymbol{\gamma}}\boldsymbol{I}\boldsymbol{R}\boldsymbol{I}^{-1}\boldsymbol{S}^{-1} = \boldsymbol{S}^{-1}\boldsymbol{I}^{-1}[\boldsymbol{R}]_{\cdot,\gamma}[\boldsymbol{I}]_{\gamma,\gamma}[\boldsymbol{\Sigma}_{\gamma}]_{\gamma,\gamma}[\boldsymbol{I}]_{\gamma,\gamma}[\boldsymbol{R}]_{\gamma,\cdot}\boldsymbol{I}^{-1}\boldsymbol{S}^{-1},
$$

where notation $[\boldsymbol{A}]_{\boldsymbol{\gamma},\boldsymbol{\gamma}}$ means the square submatrix of $\boldsymbol{A}$ corresponding to the indexes in $\Gamma_{\boldsymbol{\gamma}}$ and the dot notation in the subscripts $(\cdot)$ means all row or column indexes. Define

$$
\boldsymbol{N} := \text{Diag}\left(\sqrt{n_j}\right),
$$

where $n_j$ is the variant specific sample size in the meta-analysis. Then

$$
\boldsymbol{S}^{-1}\boldsymbol{I}^{-1} \approx \boldsymbol{N}
$$

In practice we observe that if there is any error in the scaling of variants or the measurement of imputation INFO scores, then substituting $\boldsymbol{S}^{-1}\boldsymbol{I}^{-1}$ with $\boldsymbol{N}$ will result in more accurate fine-mapping. In future derivations we replace $\boldsymbol{S}^{-1}\boldsymbol{I}^{-1}$ with $\boldsymbol{N}$. To further simplify notation, define a $k_\gamma \times k_\gamma$ matrix

$$
\boldsymbol{Q}_{\boldsymbol{\gamma}} := \boldsymbol{N}[\boldsymbol{R}]_{\cdot,\gamma}[\boldsymbol{I}]_{\gamma,\gamma}[\boldsymbol{\Sigma}_{\gamma}^{\frac{1}{2}}]_{\gamma,\gamma} \in \mathbb{R}^{p \times k},
$$

so that

$$
L(\boldsymbol{\gamma}; \text{DATA}) \propto \mathcal{N}(\widehat{\boldsymbol{z}}|\mathbf{0}_p, \boldsymbol{R}_{\boldsymbol{M}} + \boldsymbol{Q}_{\boldsymbol{\gamma}}\boldsymbol{Q}_{\boldsymbol{\gamma}}^{\intercal}).
$$

This form highlights that the likelihood of a causal configuration can be obtained by performing a rank-$k_\gamma$ update on the covariance matrix of the null configuration, which enables efficient methods to perform the required matrix computations when $k_{\boldsymbol{\gamma}}$ is small, i.e., when the causal configuration is sparse.

### 3 Efficient implementation

#### 3.1 Matrix inversion and determinants

FINEMAP-miss evaluates the relative evidence of a configuration $\boldsymbol{\gamma}$ through its logarithmic Bayes factor, comparing the marginal likelihood of $\boldsymbol{\gamma}$ to that of the null configuration $\boldsymbol{\gamma}_0 := \{0\}^p$.

$$
\begin{aligned}
\log\big(\text{BF}_{\boldsymbol{\gamma}}\big) &= \log\big(L(\boldsymbol{\gamma}; \text{DATA})\big) - \log\big(L(\boldsymbol{\gamma}_0; \text{DATA})\big) \\
&= -\frac{1}{2}\Big[p \cdot \log(2\pi) + \log(|\boldsymbol{R}_{\boldsymbol{M}} + \boldsymbol{Q}_{\boldsymbol{\gamma}}\boldsymbol{Q}_{\boldsymbol{\gamma}}^{\intercal}|) + \widehat{\boldsymbol{z}}^{\intercal}\left(\boldsymbol{R}_{\boldsymbol{M}} + \boldsymbol{Q}_{\boldsymbol{\gamma}}\boldsymbol{Q}_{\boldsymbol{\gamma}}^{\intercal}\right)^{-1}\widehat{\boldsymbol{z}}\Big] \\
&\quad + \frac{1}{2}\left[p \cdot \log(2\pi) + \log(|\boldsymbol{R}_{\boldsymbol{M}}|) + \widehat{\boldsymbol{z}}^{\intercal}\boldsymbol{R}_{\boldsymbol{M}}^{-1}\widehat{\boldsymbol{z}}\right],
\end{aligned} \tag{11}
$$

The computational bottleneck of evaluating the log of the Bayes factor comes from the inverses and determinants of the $p \times p$ matrices $\boldsymbol{R_M}$ and $\boldsymbol{R_M} + \boldsymbol{Q_\gamma Q_\gamma^\intercal}$. The inverse and determinant of the latter matrix can be expanded using the Woodbury matrix identity, and matrix determinant lemma, respectively, as follows:

$$\left(\boldsymbol{R_M} + \boldsymbol{Q_\gamma Q_\gamma^\intercal}\right)^{-1} = \boldsymbol{R_M^{-1}} - \boldsymbol{R_M^{-1} Q_\gamma} \left(\mathbb{I}_{|\gamma|} + \boldsymbol{Q_\gamma^\intercal R_M^{-1} Q_\gamma}\right)^{-1} \boldsymbol{R_M^{-1} Q_\gamma^\intercal} \text{ and}$$
$$\left|\boldsymbol{R_M} + \boldsymbol{Q_\gamma Q_\gamma^\intercal}\right| = |\boldsymbol{R_M}| \cdot |\mathbb{I}_{|\gamma|} + \boldsymbol{Q_\gamma^\intercal R_M^{-1} Q_\gamma}|.$$

Using these rank-$k_{\boldsymbol{\gamma}}$ matrix updates, the log of the Bayes factor in equation (11) can be updated into a more efficient form

$$\begin{aligned}
\log(\mathrm{BF}_{\boldsymbol{\gamma}}) = &-\frac{1}{2}\left[p \cdot \log(2\pi) + \log(|\boldsymbol{R_M}|) + \log(|\mathbb{I}_{|\gamma|} + \boldsymbol{Q_\gamma^\intercal R_M^{-1} Q_\gamma}|)\right. \\
&\quad \left. + \widehat{\boldsymbol{z}}^\intercal \left(\boldsymbol{R_M^{-1}} - \boldsymbol{R_M^{-1} Q_\gamma} (\mathbb{I}_{|\gamma|} + \boldsymbol{Q_\gamma^\intercal R_M^{-1} Q_\gamma})^{-1} \boldsymbol{Q_\gamma^\intercal R_M^{-1}}\right) \widehat{\boldsymbol{z}}\right] \\
&+ \frac{1}{2}\left[p \cdot \log(2\pi) + \log(|\boldsymbol{R_M}|) + \widehat{\boldsymbol{z}}^\intercal \boldsymbol{R_M^{-1}} \widehat{\boldsymbol{z}}\right] \\
= &-\frac{1}{2}\left[\log(|\mathbb{I}_{|\gamma|} + \boldsymbol{Q_\gamma^\intercal R_M^{-1} Q_\gamma}|)\right. \\
&\quad \left. - \widehat{\boldsymbol{z}}^\intercal \boldsymbol{R_M^{-1} Q_\gamma} (\mathbb{I}_{|\gamma|} + \boldsymbol{Q_\gamma^\intercal R_M^{-1} Q_\gamma})^{-1} \boldsymbol{Q_\gamma^\intercal R_M^{-1}} \widehat{\boldsymbol{z}}\right]. \qquad (12)
\end{aligned}$$

Since $\boldsymbol{R_M^{-1}}$ does not depend on $\boldsymbol{\gamma}$, it needs to be evaluated only once for the entire analysis, while $\mathbb{I}_{|\gamma|} + \boldsymbol{Q_\gamma^\intercal R_M^{-1} Q_\gamma}$ is a $k_{\boldsymbol{\gamma}} \times k_{\boldsymbol{\gamma}}$ matrix, whose inverse and determinant can be quickly computed when $\boldsymbol{\gamma}$ is a sparse causal configuration for which $k_{\boldsymbol{\gamma}} \ll p$.

#### 3.2 Inverting nearly singular matrices

By using the Woodbury matrix identity there is no longer a need to invert a $p \times p$ matrix for every configuration, but one inversion of $\boldsymbol{R_M}$ is still required. In practice, this cannot always be done without some modifications to $\boldsymbol{R_M}$, since it can be numerically unstable when some pairs of variants are highly correlated.

Diagonal adjustment methods have been considered by Zou et al. [4], Schäfer et al. [5], and Wu et al. [6] to resolve issues arising from singular matrices. Following this idea, we choose a constant $\varepsilon > 0$ and shrink the off-diagonal terms of $\boldsymbol{R_M}$ by multiplying them with $1 - \varepsilon$ while the diagonal will remain as 1:

$$\boldsymbol{R_M^\varepsilon} := (1 - \varepsilon) \cdot \boldsymbol{R_M} + \varepsilon \cdot \mathbb{I}_p.$$

To simplify notation, we will assume by default that $\boldsymbol{R_M}$ has been adjusted appropriately so that it is invertible in a numerically stable way, and we omit the $\varepsilon$ superscript.

#### 3.3 Pre-multiplication

FINEMAP-miss is further optimized by pre-multiplying matrices to avoid redundant computations. Let $\boldsymbol{\gamma}_1 := \{1\}^p$ denote the saturated configuration, and consider the vector

$$\boldsymbol{v} := \widehat{\boldsymbol{z}}^\intercal \boldsymbol{R_M^{-1} Q_{\gamma_1}}.$$

For any given configuration $\boldsymbol{\gamma}$, we can obtain $\widehat{\boldsymbol{z}}^\intercal \boldsymbol{R_M^{-1} Q_\gamma}$ by extracting the elements from $\boldsymbol{v}$ corresponding to the causal variants in $\boldsymbol{\gamma}$,

$$\widehat{\boldsymbol{z}}^\intercal \boldsymbol{R_M^{-1} Q_\gamma} = [\boldsymbol{v}]_{\boldsymbol{\gamma}}.$$

A similar pre-computation can be performed for the determinant since if we define

$$\boldsymbol{D} := \mathbb{I}_p + \boldsymbol{Q_{\gamma_1}^T R_M^{-1} Q_{\gamma_1}},$$

then

$$\mathbb{I}_{|\gamma|} + \boldsymbol{Q_\gamma^T R_M^{-1} Q_\gamma} = [\boldsymbol{D}]_{\boldsymbol{\gamma},\boldsymbol{\gamma}}.$$

The computational complexity of $\boldsymbol{v}$ and $\boldsymbol{D}$ are $\mathcal{O}(p^3)$. These are higher compared to directly computing $\widehat{\boldsymbol{z}}^\intercal \boldsymbol{R_M^{-1} Q_\gamma}$ and $\mathbb{I}_{|\gamma|} + \boldsymbol{Q_\gamma^T R_M^{-1} Q_\gamma}$ for any one $\boldsymbol{\gamma}$, which have a complexity $\mathcal{O}(kp^2)$, but since hundreds of thousands of configurations may need to be evaluated, these pre-computations save time in the long run.

#### 4 Avoiding local maxima with multiple causal variants

When running FINEMAP-miss on simulated data sets containing multiple causal variants, we observed scenarios where the Shotgun Stochastic Search (SSS) [7] algorithm got stuck in local maxima, and the resulting PIPs from fine-mapping did not reveal the true causal variants. This happens when the LD structure makes the marginal effects of the causal variants cancel out, while simultaneously the marginal effect of some non-causal variant becomes magnified. This problem is exacerbated when the causal variants are missing data compared to the non-causal one, since then the SSS algorithm may get stuck around configurations containing the non-causal, fully observed variant. This is possible since the SSS only adds, removes or changes one variant at a time, and hence may miss the configurations that contain both causal variants, even when those configurations would have the highest log BFs.

To help FINEMAP-miss find causal configurations with high log BFs, we developed an algorithm to find a suitable starting point for SSS.

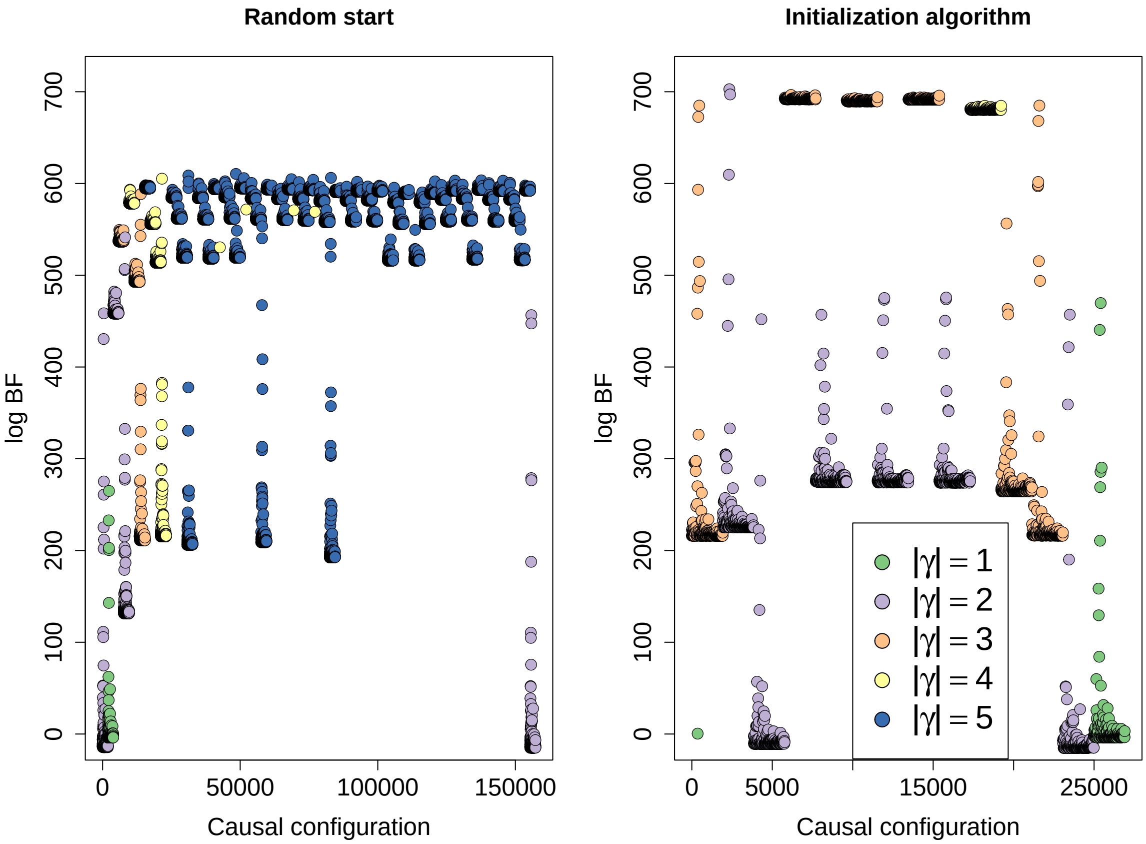


Figure 1: Two runs of FINEMAP-miss of a simulated GWAS meta-analysis with $p = 1925$ variants, of which two were causal. The plots display the log Bayes Factors (BF) of the causal configurations in the order that they were evaluated. In the left plot, FINEMAP-miss was initialized with a random configuration of size $|\gamma| = 1$. In the right plot, the new initialization algorithm was used with $n_e = 10{,}000$, $|\gamma^e| = 100$. $|\gamma| = 2$ was used for the initial configuration. Colors indicate the size of the causal configuration as given in the legend.

In figure 1, we demonstrate that FINEMAP-miss can fail to evaluate the correct causal configuration when initialized with a random starting configuration (left), as the algorithm begins searching for increasingly larger configurations, that do not contain the true causal variants (max. log-BF $\approx 611$). When using our initialization algorithm to find a better start (right), SSS is able to quickly evaluate configurations containing the true causal variants (max. log-BF $\approx 703$).

The initialization algorithm works by exploring a set of exploratory configurations $\gamma^e$ that are large (e.g. $|\gamma^e| = 100$). For the algorithm, choose $n_e$, the number of exploratory configurations to be checked. Then,

1. For $i = 1, \dots, n_e$ :
   (a) Create an exploratory configuration $\gamma_i^e$ by sampling $|\gamma^e|$ variants at random.
   (b) Evaluate the log BF of $\gamma_i^e$.
2. Select the top decile of the exploratory configurations by log BFs.
3. Create the start configuration of size $|\gamma| = k$ by choosing the $k$ variants that are most frequently shared between the top exploratory configuration (having the highest BF) and the other exploratory configurations of the top decile.

The rationale for this algorithm follows from the idea that any configuration containing the true causal variants will have a higher log BF compared to configurations in which they are not present. For suitably large $|\gamma^e|$, and $n_e$, we can be quite certain that at least one exploratory configuration will contain a combination of the true causal variants.

To quantify the success probability of the algorithm, let $X$ be a random variable describing the number of causal variants in $\gamma^e$ and suppose there truly are $c$ causal variants among the set of $p$ variants. Then the probability of sampling all causal variants in an exploratory configuration with size $|\gamma^e|$ can be derived from a hypergeometric distribution.

$$\mathbb{P}(X = c) = \frac{\binom{c}{c}\binom{p-c}{|\gamma^e|-c}}{\binom{p}{|\gamma^e|}} = \frac{\binom{p-c}{|\gamma^e|-c}}{\binom{p}{|\gamma^e|}}.$$

If we check $n_e$ exploratory configurations, the probability of sampling all causal variants at least once in some $\gamma_i^e$ is

$$1 - [1 - \mathbb{P}(X = c)]^{n_e} .$$

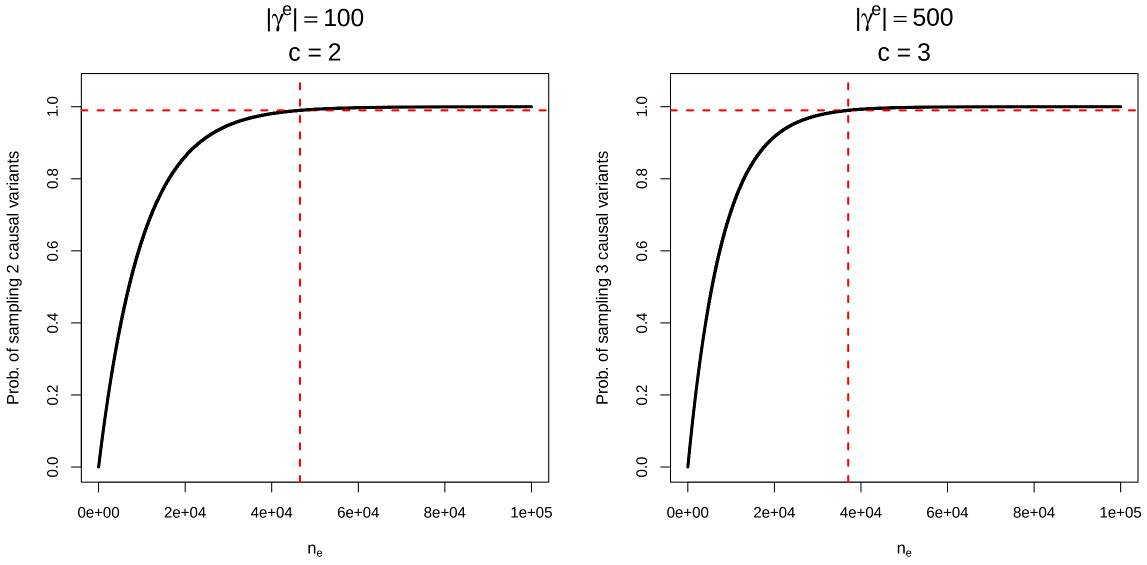


Figure 2: Probability of sampling all causal variants at least once in an exploratory configuration as a function of $n_e$ for given values of $|\gamma^e|$, when $p$ = 10,000. The red dashed lines indicate the minimum value of $n_e$ for which the probability of observing the causal variants is above 0.99.

By varying $n_e$ and $|\gamma^e|$, we can control the probability that a combination of the true causal variants are observed in at least one $\gamma_i^e$. For example, with $c = 2$, $|\gamma^e| = 100$, setting $n_e = 46,510$ is sufficient to have a probability above 0.99 to sample both causal variants at least once. For $c = 3$, $|\gamma^e| = 500$, setting $n_e = 37,051$ is sufficient. Here, we do need to note that as the parameters $c$, $n_e$, $|\gamma^e|$, and $p$ increase, the time it takes to find the starting configuration also increases, since each $\gamma_i^e$ requires an inversion of a $|\gamma^e| \times |\gamma^e|$ matrix.

The algorithm considers only variants included in the top configuration rather than choosing the $k$ most frequent variants among all configurations in the top decile. This is because we are interested in finding some combination of variants that jointly explain the data very well, whereas the most frequent $k$ variants in the top decile could be a set of variants that are highly correlated to only one of the causal variants.

In principle, starting fine-mapping with an initial configuration of size greater than 1 could result in some singleton configurations not being evaluated. In the implementation of FINEMAP-miss, we have ensured that all singleton configurations are automatically evaluated.

We note that the search for an optimal starting configuration is not guaranteed to avoid problems with local maxima, since it may be difficult to choose an appropriate size for the initial configuration, and there still remains a small probability that an optimal combination of variants has not been found in the search.

### 5 Posterior inclusion probability (PIP) bins

In simulated data, the PIP values can be used for benchmarking the performance of fine-mapping methods [8]. For a correclty calibrated fine-mapping model, the causal status $\gamma_j \in \{0, 1\}$ of variant $j$ should be described well by the Bernoulli distribution

$$\gamma_j \sim \text{Bernoulli}(\text{PIP}_j),$$

where $\text{PIP}_j$ is the PIP given by the model to variant $j$. It follows that for any set $J$ of variants, a correctly calibrated model would have the property that

$$\frac{1}{|J|}\mathbb{E}\left[\sum_{j\in J}\gamma_j\right] = \frac{1}{|J|}\sum_{j\in J}\text{PIP}_j.$$

In particular, the PIPs should have the property that among all variants with PIP equal to $P$, the proportion $P$ truly are causal. It is difficult to assess the model performance for a single value of PIP, so we group variants with similar PIPs into bins. We split the range [0,1] into $I + 1$ intervals $[0, t_1)$, $[t_1, t_2)$, $\ldots$ $[t_{I-1}, t_I)$, $[t_I, 1]$, where $t_1, \ldots, t_I$ are fixed cut points, and assign each variant to the interval corresponding to its PIP. Denote the $i$th PIP bin by $b_i$ and define $\overline{\text{PIP}}_i := \frac{1}{|b_i|}\sum_{j\in b_i}\text{PIP}_j$. If the PIPs are well calibrated, we expect the proportion of causal variants in the bin to be close to the average PIP of the bin,

$$\frac{1}{|b_i|}\sum_{j\in b_i}\gamma_j \approx \overline{\text{PIP}}_i.$$

To obtain reasonable confidence intervals for finite samples, we consider the causal status of variants from a single bin $b_i$ to come from a binomial distribution,

$$\sum_{j\in b_i}\gamma_j \sim \text{Bin}\left(|b_i|, \overline{\text{PIP}}_i\right).$$

For the confidence interval, we use the Wilson score interval, which is more accurate than the Normal approximation for bins close to the end points (0 or 1). The interval for bin $b_i$ is given as

$$\frac{1}{1 + 1.96^2/|b_i|}\left(\overline{\text{PIP}}_i \pm \frac{1.96}{2|b_i|}\sqrt{4|b_i|\overline{\text{PIP}}_i(1 - \overline{\text{PIP}}_i) + 1.96^2}\right).$$

If the proportion of causal variants in PIP bins lies outside the confidence interval more often than expected by chance, then we conclude that the fine-mapping method is likely miscalibrated.

### 6 Breast cancer meta-analysis details

The summary statistics taken from a study by Rashkin et al. [9] are a meta-analysis of a case-control GWAS for breast cancer that combined UK biobank and GERA datasets. The summary data available through the GWAS catalog consisted of p-values and odds-ratios for each analyzed variant. To obtain z-scores for fine-mapping, for variant $j$, the p-value, $p_j$, was transformed using the inverse-normal cumulative density function $\Phi^{-1} : [0,1] \to \mathbb{R}_{\geq 0}$ to yield the absolute value of the z-score

$$|\widehat{z}_j| = \Phi^{-1}\left(1 - \frac{p_j}{2}\right).$$

To obtain the correct sign for the z-score, we take the sign from the log odds ratio.

$$\operatorname{sgn}(\widehat{z}_j) = \operatorname{sgn}\left(\log\left(\mathrm{OR}_j\right)\right).$$

Effective sample sizes were obtained from the counts of cases and controls per cohort, using the formula $n_{\text{eff}} = \frac{n_{\text{case}} \times n_{\text{control}}}{n_{\text{case}} + n_{\text{control}}}$.

| Cohort | Cases | Controls | Effective sample size |
|---|---|---|---|
| UKB | 13903 | 189855 | $N_U = 12954$ |
| GERA | 3978 | 29801 | $N_G = 3510$ |

From the effective sample sizes, we can estimate the standard errors of the meta-analyzed standardized marginal effects.

$$s_{j,\text{Meta}} \approx \left[N_{\text{UKB}} \cdot \mathcal{I}_{\text{UKB}}(j) + N_{\text{GERA}} \cdot \mathcal{I}_{\text{GERA}}(j)\right]^{-\frac{1}{2}},$$

where $\mathcal{I}_{\text{UKB}}, \mathcal{I}_{\text{GERA}}$ are indicator functions, measuring whether the variant $j$ was observed in UKB and GERA, respectively.

Since the data was provided in a meta-analyzed form, we were not able to differentiate the marginal effects between UKB and GERA, except in the cases when the variant had only been observed in one of the studies in which case the meta-analyzed value was equal to the estimate from that study. This lack of information about the effects between cohorts impedes accurate summary statistic imputation. However, we still attempted an approximate imputation by assuming that the effects were the same in both studies. The marginal effects are then obtained by multiplying the meta-analysis z-score by the corresponding standard error and assigning this same value to both individual cohorts.

$$\widehat{\beta}_{j,\text{UKB}} = \widehat{\beta}_{j,\text{GERA}} = \widehat{\beta}_{j,\text{Meta}} = \widehat{z}_j \cdot s_{j,\text{Meta}}.$$

The cohort based standard errors, whenever the variant was observed, were approximated using the effective sample sizes.

$$s_{j,\text{UKB}} \approx {N_U}^{-\frac{1}{2}},$$
$$s_{j,\text{GERA}} \approx {N_G}^{-\frac{1}{2}}.$$

### 7 Posterior expected number of causal variants (PENC)

In the main text, we defined PENC statistic and compared it between the methods for the case of simulations with one causal variant. The distributions of PENC are visualized in Supplementary Figures 3 and 4 for high and low INFO causal variants, respectively.

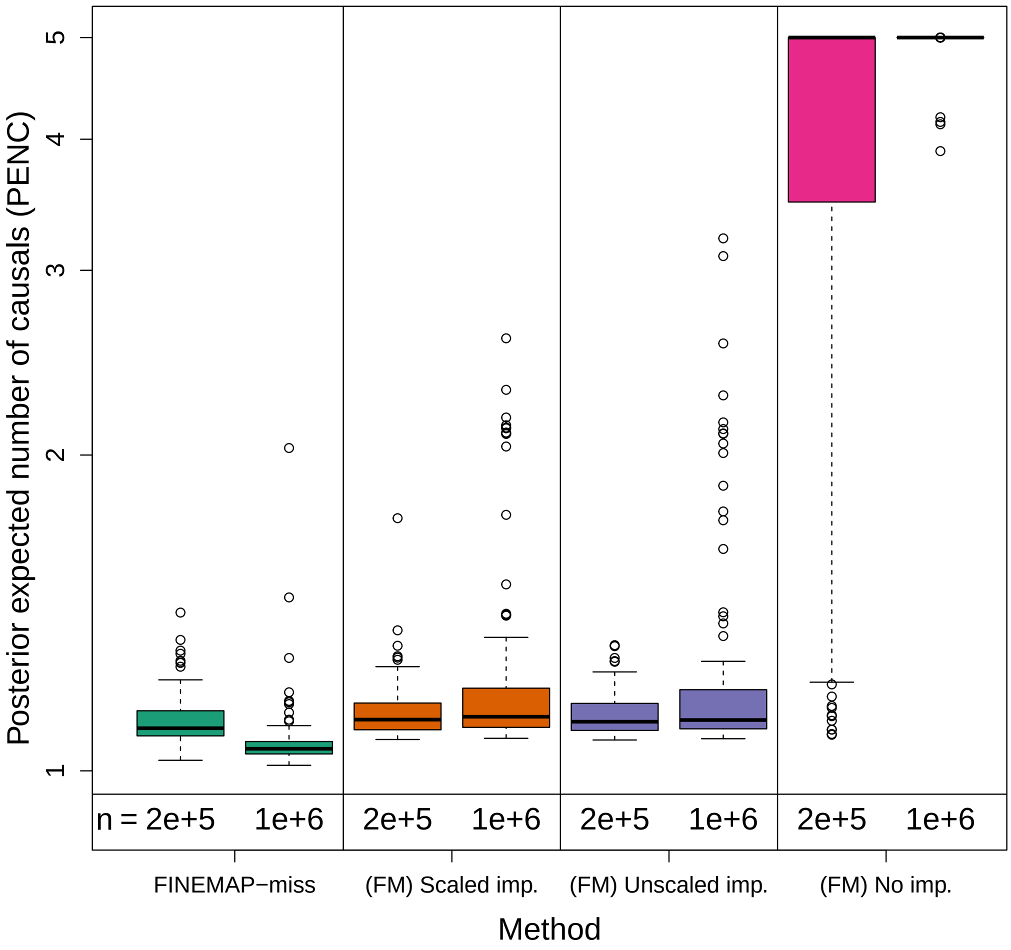


Figure 3: PENC from the meta-analysis fine-mapping simulations with one causal variant that had a high imputation INFO ($> 0.90$). The simulations are separated by sample sizes $n$ = 2e+5 or $n$ = 2e+5. The four methods are FINEMAP-miss v.1.0 and FINEMAP v.1.4.2 that was run with three versions of the input data: scaled summary statistics imputation, unscaled summary statistics imputation or no summary statistics imputation. FM = FINEMAP v.1.4.2. imp. = imputed data.

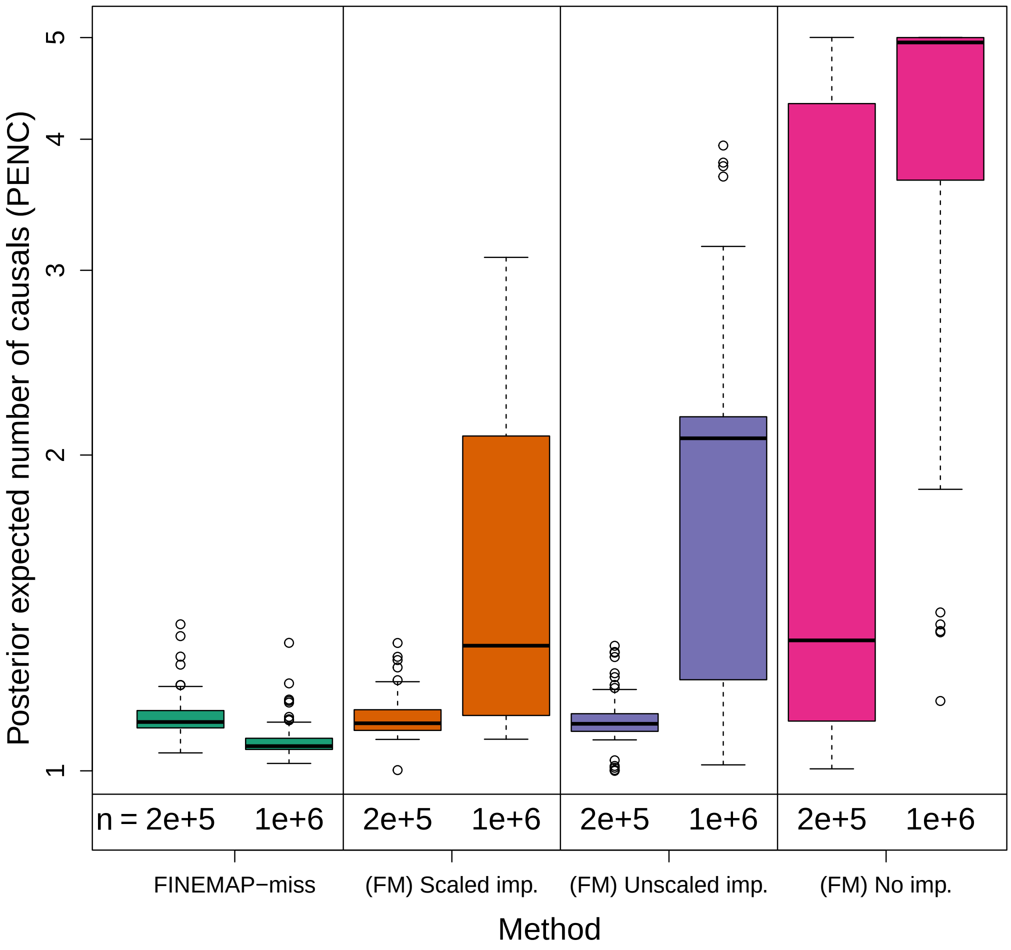


Figure 4: PENC from the meta-analysis fine-mapping simulations with one causal variant that had a low imputation INFO ($< 0.90$). The simulations are separated by sample sizes $n$ = 2e+5 or $n$ = 1e+6. The four methods are FINEMAP-miss v.1.0 and FINEMAP v.1.4.2 that was run with three versions of the input data: scaled summary statistics imputation, unscaled summary statistics imputation or no summary statistics imputation. FM = FINEMAP v.1.4.2. imp. = imputed data.

### 8 Simulations with two causal variants

This section reports the fine-mapping results from simulations with two causal variants in Supplementary Table 1 and Supplementary Figures 3,4 and 5. In each simulation, each causal variant had a high INFO ($> 0.9$), and explained 0.1% of phenotypic variance.

Table 1: Means of credible set (CS) power, PIP of causal variant (C-PIP), maximal PIP of non-causal variants (MNC-PIP) and PENC for from meta-analysis fine-mapping simulations with 2 causal variants (INFO $> 0.9$), separated by sample size $n$ = 2e+5 or $n$ = 1e+6. FINEMAP v.1.4.2 was run with three versions of the input data: scaled imputation, unscaled imputation or no imputation. FM = FINEMAP v.1.4.2. imp. = imputed data.

| | Method | | | |
|---|---|---|---|---|
| | FINEMAP-miss | (FM) Scaled imp. | (FM) Unscaled imp. | (FM) No imp. |
| | | $n$ = 2e+5 | | |
| CS power | 0.990 | 0.980 | 0.975 | 0.160 |
| C-PIP | 0.476 | 0.486 | 0.460 | 0.132 |
| MNC-PIP | 0.270 | 0.248 | 0.277 | 0.988 |
| PENC | 2.118 | 2.132 | 2.132 | 4.852 |
| | | $n$ = 1e+6 | | |
| CS power | 0.995 | 0.985 | 0.885 | 0.145 |
| C-PIP | 0.576 | 0.573 | 0.450 | 0.145 |
| MNC-PIP | 0.312 | 0.404 | 0.572 | 1.000 |
| PENC | 2.082 | 2.313 | 2.441 | 5.000 |

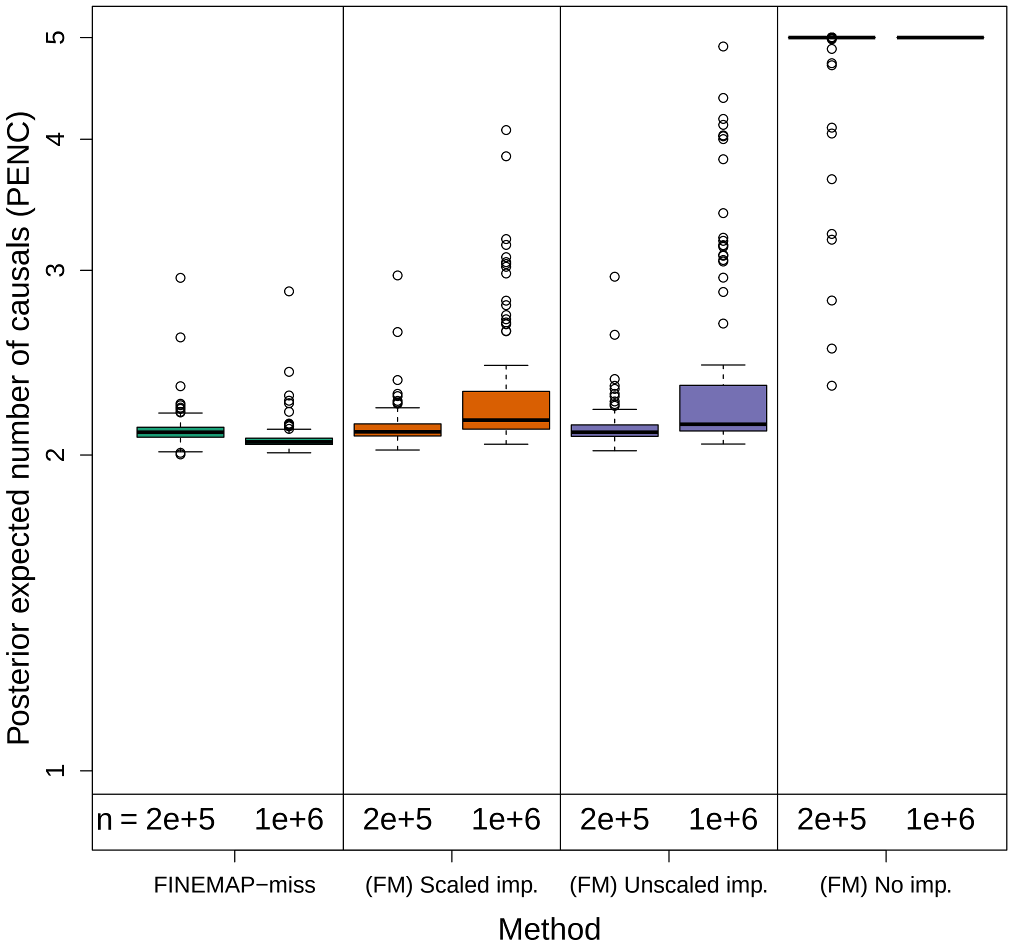


Figure 5: PENC from the meta-analysis fine-mapping simulations with two causal variants that both had a high imputation INFO (> 0.90). The simulations are separated by sample sizes $n$ = 2e+5 or $n$ = 1e+6. The four methods are FINEMAP-miss v.1.0 and FINEMAP v.1.4.2 that was run with three versions of the input data: scaled summary statistics imputation, unscaled summary statistics imputation or no summary statistics imputation. FM = FINEMAP v.1.4.2. imp. = imputed data.

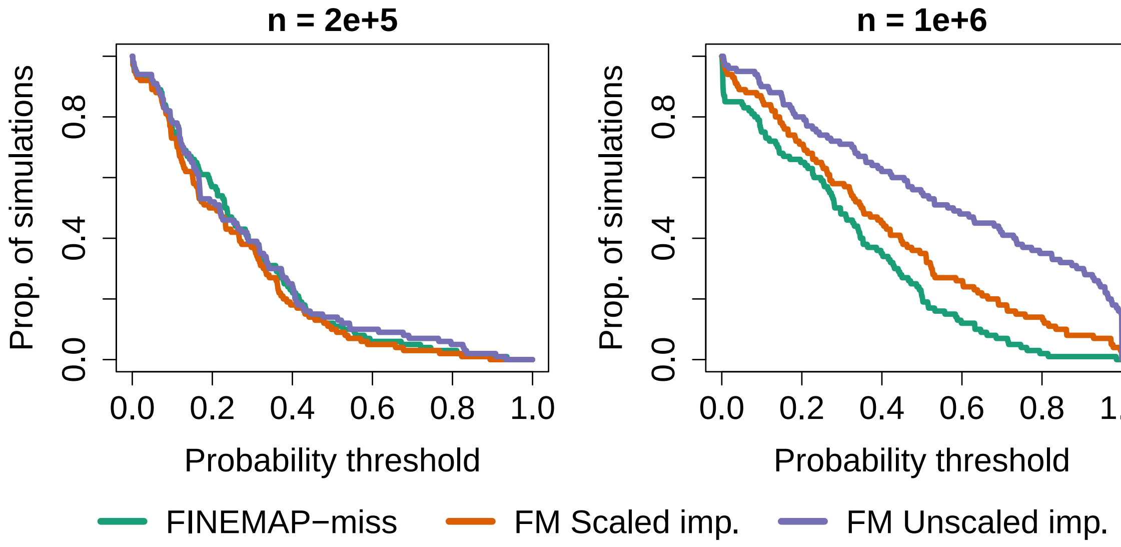


Figure 6: Proportion of simulations (y-axis), whose fine-mapping results contained a non-causal variant with a PIP above a probability threshold (x-axis). Results displayed separately for simulations with sample sizes $n$ = 2e+5 and $n$ = 1e+6. The simulations had 2 causal variants with high INFO ($> 0.9$). The three methods are FINEMAP-miss v.1.0 and FINEMAP v.1.4.2 that was run with two versions of the input data: scaled summary statistics imputation or unscaled summary statistics imputation. FM = FINEMAP v.1.4.2. imp. = imputed data.

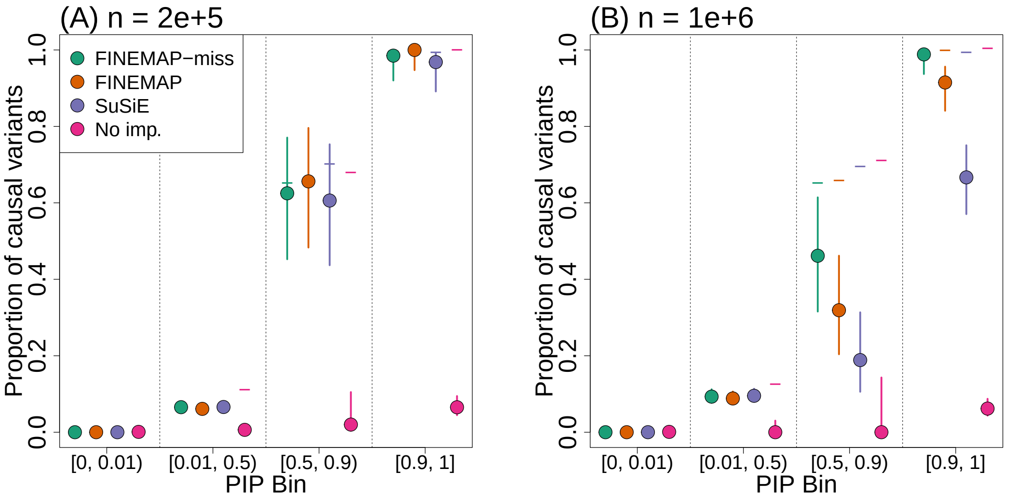


Figure 7: PIP bin calibration plot from simulations with two causal variants with high INFO ($> 0.90$). Points indicate the observed proportion of causal variants. Horizontal lines indicate the expectation given by the average PIPs. 95% Wilson confidence intervals are provided as vertical lines. The simulation sample size is 2e+5 in panel A and $n$ = 1e+6 in panel B. FM = FINEMAP v.1.4.2. imp. = imputed data.FM = FINEMAP v.1.4.2. imp. = imputed data. For the smaller sample size, the PIP bins are well calibrated for FINEMAP-miss and FINEMAP with scaled imputed summary statistics. For $n$ = 1e+6, both methods possibly exhibit some miscalibration in a middle bin $[0.5, 0.9)$. FINEMAP-miss is the only method, for which the top bin is well calibrated.

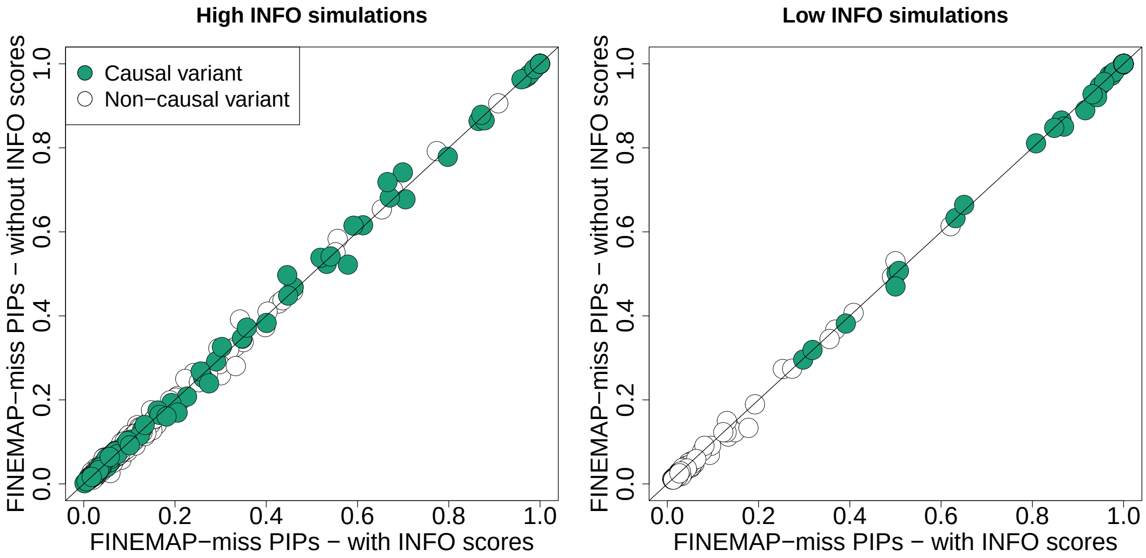


Figure 8: Comparison of the PIPs from FINEMAP-miss, when INFO scores were provided as input (x-axis) and when they were set to 1 (y-axis). Variants with both PIPs below 0.01 are omitted from the plot. Data are from simulations with one causal variant that had either a high INFO (>0.9) (left panel) or low info (<0.9) (right panel).

### 9 INFO scores have little impact on the output of FINEMAP-miss

With FINEMAP-miss, we have derived a fine-mapping model for situation where each variant has its own imputation INFO value that reflects how accurately we know the genotype values / GWAS summary statistics at the variant. Thus, we investigated the possible effect that changes in the INFO scores have on the performance of FINEMAP-miss. For this, we redid the fine-mapping of the simulations with one causal variant with FINEMAP-miss, but instead of providing the true INFO scores to the model, we set each INFO score to be 1, which ignores any variation in INFO scores between the variants.

A comparison of the fine-mapping output is provided in Supplementary Figure 8, which demonstrates that the variant-specific PIPs are nearly identical between the two versions of the INFO scores given as input. This suggests that accounting for the INFO scores within the FINEMAP model does not make a noticeable difference. Intuitively, we assume that this is because the effect of INFO scores is already carried by the z-scores, that account for the estimated effects and increased standard errors due to the imputed dosage data, which appears to be enough for comparing the possible causality of the variants relative to each other. In the next section, we provide also an analytical explanation for this behavior.

If inclusion of the INFO scores does not affect the performance of the FINEMAP model, why FINEMAP-miss then performed better than FINEMAP with imputed summary statistics, especially when causal variant had a low INFO? We expect that this is simply because summary statistics imputation quality is poor for low INFO variants, which biases the fine-mapping of imputed data and this bias becomes apparent when the causal variants have low INFO.

### 10 The impact of INFO scores on Bayes factors with growing sample size

Consider $\boldsymbol{Q_\gamma} = \boldsymbol{S}^{-1}\boldsymbol{I}^{-1}[\boldsymbol{R}]_{\cdot,\gamma}[\boldsymbol{I}]_{\gamma,\gamma}[\boldsymbol{\Sigma_\gamma}]^{\frac{1}{2}}_{\gamma,\gamma}$ and define the matrix

$$\boldsymbol{Q}^*_{\boldsymbol{\gamma}} := [\boldsymbol{R}]_{\cdot,\gamma}[\boldsymbol{\Sigma_\gamma}]^{\frac{1}{2}}_{\gamma,\gamma} = \boldsymbol{N}^{-1}\boldsymbol{Q_\gamma}\boldsymbol{I}^{-1}_{\boldsymbol{\gamma}} \quad \text{so that} \quad \boldsymbol{Q_\gamma} = \boldsymbol{N}\boldsymbol{Q}^*_{\boldsymbol{\gamma}}\boldsymbol{I_\gamma},$$

where $\boldsymbol{I}_{\boldsymbol{\gamma}} := [\boldsymbol{I}]_{\boldsymbol{\gamma},\boldsymbol{\gamma}}$. By substituting this form in the place of $\boldsymbol{Q}_{\boldsymbol{\gamma}}$ in the log Bayes factor formula (12),

$$\log(\mathrm{BF}_{\boldsymbol{\gamma}}) = -\frac{1}{2}\left[\log(|\mathbb{I}_{|\gamma|} + \boldsymbol{I}_{\gamma}\boldsymbol{Q}^{*\intercal}_{\gamma}\boldsymbol{N}\boldsymbol{R}^{-1}_{\boldsymbol{M}}\boldsymbol{N}\boldsymbol{Q}^{*}_{\gamma}\boldsymbol{I}_{\gamma}|) \right. \\ \left. -\widehat{\boldsymbol{z}}^{\intercal}\boldsymbol{R}^{-1}_{\boldsymbol{M}}\boldsymbol{N}\boldsymbol{Q}^{*}_{\gamma}\boldsymbol{I}_{\gamma}(\mathbb{I}_{|\gamma|} + \boldsymbol{I}_{\gamma}\boldsymbol{Q}^{*\intercal}_{\gamma}\boldsymbol{N}\boldsymbol{R}^{-1}_{\boldsymbol{M}}\boldsymbol{N}\boldsymbol{Q}^{*}_{\gamma}\boldsymbol{I}_{\gamma})^{-1}\boldsymbol{I}_{\gamma}\boldsymbol{Q}^{*\intercal}_{\gamma}\boldsymbol{N}\boldsymbol{R}^{-1}_{\boldsymbol{M}}\widehat{\boldsymbol{z}}\right]. \tag{13}$$

As the variant sample sizes increase, the impact of the constant identity matrix $\mathbb{I}_{|\gamma|}$ becomes negligible in the log Bayes factor compared to the term involving $\boldsymbol{N}$, so that

$$\begin{aligned}\log(\mathrm{BF}_{\boldsymbol{\gamma}}) &\approx -\frac{1}{2}\left[\log(|\boldsymbol{I}_{\gamma}\boldsymbol{Q}^{*\intercal}_{\gamma}\boldsymbol{N}\boldsymbol{R}^{-1}_{\boldsymbol{M}}\boldsymbol{N}\boldsymbol{Q}^{*}_{\gamma}\boldsymbol{I}_{\gamma}|) \right.\\ &\qquad \left. -\widehat{\boldsymbol{z}}^{\intercal}\boldsymbol{R}^{-1}_{\boldsymbol{M}}\boldsymbol{N}\boldsymbol{Q}^{*}_{\gamma}\boldsymbol{I}_{\gamma}(\boldsymbol{I}_{\gamma}\boldsymbol{Q}^{*\intercal}_{\gamma}\boldsymbol{N}\boldsymbol{R}^{-1}_{\boldsymbol{M}}\boldsymbol{N}\boldsymbol{Q}^{*}_{\gamma}\boldsymbol{I}_{\gamma})^{-1}\boldsymbol{I}_{\gamma}\boldsymbol{Q}^{*\intercal}_{\gamma}\boldsymbol{N}\boldsymbol{R}^{-1}_{\boldsymbol{M}}\widehat{\boldsymbol{z}}\right]\\ &= -\frac{1}{2}\left[\log(|\boldsymbol{I}_{\gamma}\boldsymbol{Q}^{*\intercal}_{\gamma}\boldsymbol{N}\boldsymbol{R}^{-1}_{\boldsymbol{M}}\boldsymbol{N}\boldsymbol{Q}^{*}_{\gamma}\boldsymbol{I}_{\gamma}|) \right.\\ &\qquad \left. -\widehat{\boldsymbol{z}}^{\intercal}\boldsymbol{R}^{-1}_{\boldsymbol{M}}\boldsymbol{N}\boldsymbol{Q}^{*}_{\gamma}(\boldsymbol{Q}^{*\intercal}_{\gamma}\boldsymbol{N}\boldsymbol{R}^{-1}_{\boldsymbol{M}}\boldsymbol{N}\boldsymbol{Q}^{*}_{\gamma})^{-1}\boldsymbol{Q}^{*\intercal}_{\gamma}\boldsymbol{N}\boldsymbol{R}^{-1}_{\boldsymbol{M}}\widehat{\boldsymbol{z}}\right]\\ &= -\frac{1}{2}\left[\log(|\boldsymbol{Q}^{*\intercal}_{\gamma}\boldsymbol{N}\boldsymbol{R}^{-1}_{\boldsymbol{M}}\boldsymbol{N}\boldsymbol{Q}^{*}_{\gamma}|) + \log(|\boldsymbol{I}_{\gamma}\boldsymbol{I}_{\gamma}|) \right.\\ &\qquad \left. -\widehat{\boldsymbol{z}}^{\intercal}\boldsymbol{R}^{-1}_{\boldsymbol{M}}\boldsymbol{N}\boldsymbol{Q}^{*}_{\gamma}(\boldsymbol{Q}^{*\intercal}_{\gamma}\boldsymbol{N}\boldsymbol{R}^{-1}_{\boldsymbol{M}}\boldsymbol{N}\boldsymbol{Q}^{*}_{\gamma})^{-1}\boldsymbol{Q}^{*\intercal}_{\gamma}\boldsymbol{N}\boldsymbol{R}^{-1}_{\boldsymbol{M}}\widehat{\boldsymbol{z}}\right]. \end{aligned} \tag{14}$$

With this approximation, the INFO scores are separated into an additional term in the log Bayes factor formula. Let $\log(\mathrm{BF}^{\boldsymbol{I}=\mathbb{I}}_{\boldsymbol{\gamma}}) = \log(\mathrm{BF}(\boldsymbol{\gamma}|\widehat{\boldsymbol{z}}, \boldsymbol{I} = \mathbb{I}))$ denote the log Bayes factor assuming the INFO scores are all 1, corresponding to the original FINEMAP model. Then,

$$\log(\mathrm{BF}^{\boldsymbol{I}=\mathbb{I}}_{\boldsymbol{\gamma}}) - \log(\mathrm{BF}_{\boldsymbol{\gamma}}) \approx \frac{1}{2}\log(|\boldsymbol{I}_{\gamma}\boldsymbol{I}_{\gamma}|),$$

which we can write in the form

$$\log(\mathrm{BF}_{\boldsymbol{\gamma}}) \approx \log(\mathrm{BF}^{\boldsymbol{I}=\mathbb{I}}_{\boldsymbol{\gamma}}) - \frac{1}{2}\sum_{j\in\Gamma_{\gamma}}\log(\mathrm{INFO}_j). \tag{15}$$

We verified this approximation empirically using the simulations where the causal variant had a low INFO by plotting in Supplementary Figure 9 the term $\log(\mathrm{BF}_{\boldsymbol{\gamma}})$ on the y-axis while the x-axis showed the right hand side of equation (15) either with or without the correction term involving the INFO values of the variants of $\boldsymbol{\gamma}$.

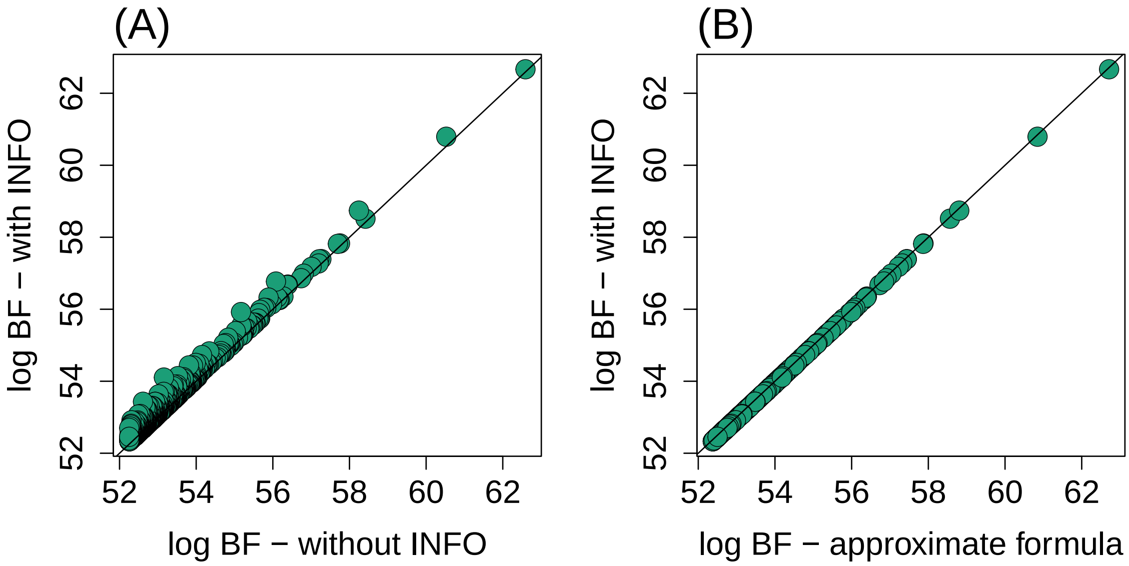


Figure 9: Effect of INFO scores on the top 1000 log Bayes factors in a single simulation ($n$ = 2e+5, causal variant has a low INFO). Y-axis shows $\log(\mathrm{BF}_{\boldsymbol{\gamma}})$, the log Bayes factors of FINEMAP-miss with true INFO scores (left side of equation (15)). X-axis of panel (A) has $\log(\mathrm{BF}^{\boldsymbol{I}=\mathbb{I}}_{\boldsymbol{\gamma}})$, the log Bayes factors from FINEMAP-miss with INFO scores set to 1, and panel (B) has the more accurate approximation using the right side of the equation (15).

In Supplementary Figure 9A, the log Bayes factors are higher for FINEMAP-miss with the true INFO scores than when INFO scores were set to 1, which is in agreement with the equation (15). In 9B, we observe that the approximation achieved by including the term $-\frac{1}{2}\sum_{j\in\Gamma_\gamma}\log(\text{INFO}_j)$ is accurate (with the given sample size $n = 2\text{e}{+}5$). We conclude that even though the INFO scores of the variants in the causal configuration affect the Bayes factor (see panel A), the effect is fairly constant across the configurations, and hence does not affect the PIPs that are computed after normalizing probabilities across the configurations. This may explain why we did not observe a noticeable difference in the PIPs depending on whether the true INFO scores were included in the analysis in Supplementary Figure 8. However, in cases where the INFO scores vary a lot across variants or where the sample size is small, a possibility to account for the exact INFO scores in fine-mapping may become more important.

#### 11 Runtime

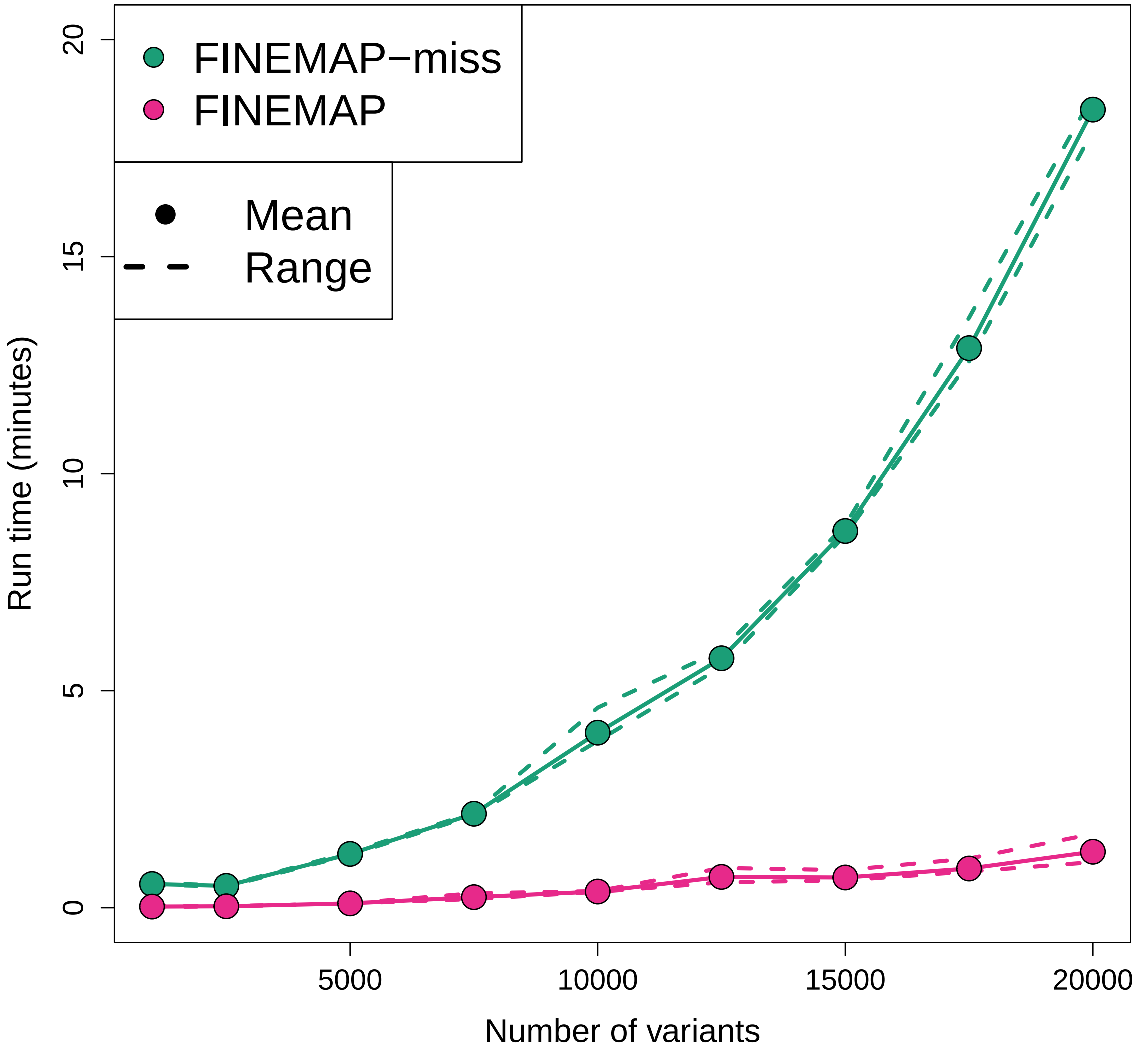


Figure 10: Run times of FINEMAP v.1.4.2 and FINEMAP-miss as a function of the number of fine-mapped variants in test runs explained in the main text.

Table 2: Run times of FINEMAP v.1.4.2 and FINEMAP-miss in minutes as a function of the number of fine-mapped variants in test runs explained in the main text.

| | **Number of variants in analysis ($p$)** | | | | | | | | |
|---|---|---|---|---|---|---|---|---|---|
| | 1,000 | 2,500 | 5,000 | 7,250 | 10,000 | 12,500 | 15,000 | 17,500 | 20,000 |
| **FINEMAP run time** | | | | | | | | | |
| min. | 0.02 | 0.03 | 0.10 | 0.20 | 0.37 | 0.58 | 0.63 | 0.83 | 1.05 |
| median | 0.01 | 0.03 | 0.10 | 0.22 | 0.37 | 0.70 | 0.65 | 0.85 | 1.08 |
| mean | 0.03 | 0.03 | 0.10 | 0.24 | 0.37 | 0.71 | 0.70 | 0.90 | 1.29 |
| max. | 0.05 | 0.03 | 0.10 | 0.33 | 0.38 | 0.92 | 0.87 | 1.13 | 1.70 |
| **FINEMAP-miss run time** | | | | | | | | | |
| min. | 0.53 | 0.49 | 1.22 | 2.13 | 3.84 | 5.54 | 8.59 | 12.59 | 17.90 |
| median | 0.54 | 0.50 | 1.23 | 2.17 | 3.92 | 5.71 | 8.61 | 12.73 | 18.44 |
| mean | 0.55 | 0.50 | 1.24 | 2.16 | 4.03 | 5.75 | 8.68 | 12.89 | 18.39 |
| max. | 0.57 | 0.51 | 1.27 | 2.19 | 4.61 | 5.93 | 8.81 | 13.59 | 18.71 |
